# DELENDA: Differentiable Epidemiology for Latent-state Estimation and Nonlinear Decision Analysis

**DOI:** 10.64898/2026.07.13.26357962

**Authors:** Joshua Suresh

## Abstract

Malaria subnational tailoring is often a population-level allocation problem: which interventions should be prioritized, at what coverage, and under what budget and uncertainty assumptions? We present Delenda, a differentiable compartmental model of *Plasmodium falciparum* transmission designed for posterior calibration and intervention-mix optimization. We fit a NUTS posterior jointly to age-stratified prevalence and clinical-incidence data from five sub-Saharan African sites plus three pre-intervention Garki Project villages, spanning a broad entomological inoculation rate (EIR) range. Delenda is implemented in JAX, which makes the full simulation differentiable. This enables efficient Bayesian inference and continuous constrained optimization over intervention coverage.

We apply the framework to an illustrative decision problem: a highly seasonal transmission setting where coverage is optimized for ITNs, SMC, IRS, and pediatric malaria vaccination across EIR, budget, objective, and uncertainty grids. Three findings are decision-relevant. First, intervention rankings are more robust than projected impact: posterior, vector-biology, and intervention-efficacy uncertainty change optimized coverage modestly but substantially widen the distribution of cases averted. Second, the objective matters: under-five optimization brings child-targeted SMC and vaccination in earlier, whereas all-age optimization delays vaccination and favors broader population protection through IRS. Third, cost uncertainty is mainly a constraint-side problem: expected-cost optima have material budget-overrun probability, while tail-risk budget rules sharply reduce overrun risk at the cost of lower effective coverage and fewer expected cases averted. Delenda therefore demonstrates an uncertainty-first approach to subnational tailoring: differentiable model structure exposes the biological parameter space to posterior calibration and carries biological and operational uncertainty into constrained decision optimization, tasks that are difficult with the non-differentiable models currently central to SNT workflows.

## 1 Introduction

Controlling malaria burden optimally reqduires effective allocation of multiple interventions, each with its own mechanism, impact, and cost. As global health financing shifts toward tighter, more explicitly prioritized budgets, programs are asked to preserve impact with less room for redundant coverage, inefficient package mixes, or poorly characterized risk. In many endemic settings, available tools—for example, insecticide-treated nets, seasonal malaria chemoprevention, indoor residual spraying, and pediatric malaria vaccines—are all plausibly useful, but their relative value depends on transmission intensity, age structure, seasonality, vector behavior, coverage history, and budget. A useful model in this context must therefore not only support counterfactual comparisons among intervention packages, but also quantify uncertainty in projected impact and expose when a recommendation is robust to the assumptions that generated it.

Mechanistic transmission models are useful precisely because transmission intensity, age-specific risk, intervention mechanism, coverage, and cost are not separable. The effect of an ITN campaign depends on mosquito survival and biting behavior; the effect of SMC or vaccination depends on the age distribution of clinical risk; the value of IRS depends on indoor resting and on how much transmission remains after nets; and all four depend on whether the setting is near elimination, in a moderate-transmission regime, or saturated by repeated infectious bites. Trial estimates and meta-analytic effect sizes are indispensable anchors, but by themselves they do not specify how effects should compose, extrapolate across EIR, or change when the objective shifts from under-five cases to all-age disease burden.

Existing malaria models span a wide range of biological and computational complexity. Ross– Macdonald-style models are transparent and analytically tractable, but they do not represent acquired immunity, age-specific clinical risk, or age-targeted intervention effects [Smith et al., 2012]. At the other extreme, individual-based simulators such as EMOD, OpenMalaria, and the Imperial malaria models encode much richer natural history and intervention mechanisms [Eckhoff, 2011, Smith et al., 2008, Griffin et al., 2010], and have been used for intervention evaluation and subnational planning [Sherrard-Smith et al., 2022, Runge et al., 2020, Ozodiegwu et al., 2023]. Those individual-based models are valuable when decisions depend on household structure, individual infection histories, local mobility, or explicit implementation processes. Many first-order subnational tailoring questions, however, pose population-level prioritization problems: which intervention class should be prioritized, at what intensity, and under what transmission, age-burden, seasonality, vector-behavior, and cost assumptions? Although individual-based models contain individual trajectories that can be interrogated, they are often validated and used at population level; the individual histories add complexity even when the decision target is an aggregate burden or allocation outcome. A structured compartmental model skips that middle step when the individual history itself is not the object of inference. For these population-level questions, simulation cost and non-differentiability reinforce each other. Calibration and optimization often expose only a subset of the biological and operational parameter space; analysts explore the remaining assumptions through scenario sweeps or local sensitivity analysis. For policy work this creates an awkward gap: the models that are easiest to optimize are often too coarse for the question, while models with more sophisticated biological representations can be hard to fit, interrogate, and use inside an uncertainty-aware optimization loop.

The bottleneck is not only simulation speed. A model used for allocation must identify which parameters the data constrain, which remain prior- or assumption-driven, and how those uncertainties propagate into the decision. A calibrated point estimate can rank packages under one set of assumptions, but it cannot distinguish a robust recommendation from one that would change under plausible alternative vector biology, intervention efficacy, or observation error. For this reason, the estimation problem and the decision problem should be coupled: the same likelihood that calibrates the transmission model should define the posterior uncertainty carried into impact projections and optimization.

This paper introduces Delenda, a model designed for this middle ground. It is a deterministic compartmental system whose human state is indexed by age, biting-risk heterogeneity, immunity, and multiplicity of infection [Cooper et al., 2019], preserving the main biological axes needed for age-targeted malaria policy without requiring individual-based simulation. This choice is not only computational: a compartmental model is parsimonious enough to make parameter identifiability visible, transparent enough to expose each intervention mechanism and prior, and fast enough to sweep transmission settings, budgets, objectives, and uncertainty sources. Delenda is implemented in JAX [Bradbury et al., 2018], making the simulation differentiable end-to-end. This lets us use gradients for likelihood-based posterior calibration and continuous constrained optimization, making high-dimensional parameter and decision spaces much more feasible to explore efficiently.

We then ask how this calibrated model can support a stylized subnational-tailoring problem. In a highly seasonal transmission scenario, we optimize continuous coverage of ITNs, SMC, IRS, and vaccination across transmission levels, budgets, and health objectives. We propagate uncertainty from four sources: the calibrated posterior, vector-biology priors, intervention-efficacy priors, and cost priors. Because vector biology and larval carrying capacity are confounded by the observed entomological inoculation rate, we condition each vector draw on the target EIR by solving for the larval capacity that reproduces the observation. Within each matched-EIR context we compare which intervention mix maximizes expected cases averted and how recommendations change when programs require greater confidence that realized costs will remain within budget.

Toward that goal, the paper contributes three pieces: a structured compartmental malaria model designed for differentiable calibration and optimization; an eight-site probabilistic calibration that exposes both well-constrained and weakly identified biological directions; and an illustrative intervention-tailoring exercise that propagates model, vector, intervention-efficacy, and cost uncertainty through competing decision criteria. The emphasis is not a deployment recommendation for a single setting, but a reusable workflow for asking which recommendations remain stable once the calibrated posterior and assumption-driven uncertainties are carried into the optimization problem.

## 2 Related Work

Malaria transmission modelling builds on the Ross–Macdonald theory of mosquito-borne infection dynamics [Smith et al., 2012], with modern policy models adding age structure, immunity, within-host infection history, vector ecology, and intervention mechanisms. OpenMalaria, EMOD, and the Imperial malaria models are the main reference points for this work [Smith et al., 2008, Eckhoff, 2011, Griffin et al., 2010]; all have informed intervention evaluation, national planning, or subnational tailoring exercises [Sherrard-Smith et al., 2022, Runge et al., 2020, Ozodiegwu et al., 2023]. Those workflows establish the biological and policy importance of mechanistic simulation, but subnational-tailoring applications typically rely on scenario sweeps, calibrated point estimates, or limited exposed parameter sets rather than full posterior propagation through continuous optimization. Delenda targets that gap: it retains a structured malaria natural history while making differentiability and likelihood-based posterior calibration first-class design goals.

Computationally, Delenda draws on differentiable scientific computing in JAX [Bradbury et al., 2018], adjoint sensitivity methods for differentiating through ODE solvers [Chen et al., 2018], automatic-differentiation variational inference [Kucukelbir et al., 2017], and NumPyro’s autoguide machinery [Phan et al., 2019].

## 3 Methods

### 3.1 Transmission model

Delenda is a coupled human–vector ODE model designed to keep the state variables most relevant for intervention prioritization: heterogeneous biting exposure, age, immunity, and multiplicity of infection in humans, coupled to a seasonal mosquito population. Most response functions are intentionally simple: infection duration, pre-erythrocytic protection, diagnostic detectability, clinical incidence, and human infectiousness are monotone or logistic functions of age, immunity, and MOI with free coefficients fit from data. This keeps the biological axes explicit while letting calibration determine the response-surface shapes. The human state is indexed by biting-risk stratum *r*, age bin *a* ∈ {1, …, *n*_*a*_} (*n*_*a*_ = 11 bins spanning [0, 75] years), immunity level *u* ∈ [0, 1] discretized on *n*_*u*_ = 10 grid points, and multiplicity of infection (MOI) *k* ∈ {0, 1, …, *K*_max_} with *K*_max_ = 15. The heterogeneous-biting analyses use five equal-weight risk strata formed from equal-probability bins of a mean-one exponential distribution, with each stratum assigned its bin mean biting multiplier. This gives most people below-average biting and a small tail with high exposure. Together these choices give a human compartmental block of dimension 5 · 11 · 10 · 16 = 8,800; the homogeneous-risk model is the special case with one risk stratum. We chose these grid sizes as a compromise between resolution and calibration speed; sensitivity checks showed that the main prevalence, incidence, and intervention rankings were not materially changed by modest refinements. We advance the ODE with an exponential-Euler update with a mass-conservation correction, rather than a plain forward-Euler step, because the infection, clearance, aging, and vector-transition rates can create large one-day compartment drains; the exponential update preserves non-negativity and is more stable while retaining a simple fixed-step solver.

Human transitions include aging across age bins, births into the youngest age bin, age-specific death rates, infection, infection clearance, and immunity movement. Demography is a closed stable population rather than a site-specific census: births enter the youngest naive, uninfected compartment at rate *bN* (*t*), individuals age between bins at the inverse bin width, and age-specific death rates use a fixed mortality-shape vector rescaled so that deaths equal births at equilibrium. We set *b* = 35.25 births per 1000 persons per year, yielding a high-fertility age pyramid roughly reflective of sub-Saharan population age structure and used for all calibration and decision simulations; the implied stable population has 14.8% under age 5 and 41.8% under age 15. In vector-driven simulations the baseline human force of infection is *λ*_0_(*t*) = *a*_eff_(*t*)*V*_*I*_ (*t*) · *V* 2*H/N*_total_(*t*); in calibration paths driven by an externally specified EIR, it is *λ*_0_(*t*) = EIR(*t*) · *V* 2*H/*365. This baseline force is distributed across biting risk, age, and immunity as

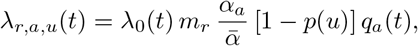

where *m*_*r*_ is the biting-risk multiplier, *α*_*a*_ is the surface-area-derived biting weight, 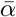 is its population average, *p*(*u*) is pre-erythrocytic protection, and *q*_*a*_(*t*) collects age-specific intervention multipliers that reduce susceptibility to new infection. New infections increment MOI by one up to *K*_max_, while clearance decrements MOI at per-brood rate *γ*(*u*), giving total clearance rate *kγ*(*u*) from MOI class *k*. Immunity increases at a rate depending on both immunity and MOI, using the fitted *b*(*u*) log(1 + *k*) form, and wanes at rate *ω*_*u*_ while uninfected. The supplementary equation table and parameter-index table define these quantities and list the fitted coefficients.

The vector subsystem tracks larval mosquitoes *V*_*L*_ and adult susceptible, exposed, and infectious mosquitoes (*V*_*S*_, *V*_*E*_, *V*_*I*_). In closed-loop vector simulations we use a linear seasonal recruitment formulation,

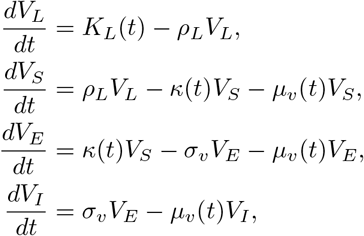

where *ρ*_*L*_ is the larval emergence rate, *σ*_*v*_ = 1*/*EIP is the sporogony rate, and *κ*(*t*) is the human-to-vector infection hazard obtained by biting-weighting infectious humans across risk, age, immunity, and MOI classes. Baseline adult biting is *a*_eff_ = *A*_base_ · ANTHR and baseline mortality is *µ*_*v*_ = *µ*_base_; interventions modify these terms by reducing successful human biting and/or increasing mosquito mortality (§3.3). The vector quantities used in the decision analysis are summarized in the supplementary vector-prior appendix. Seasonal recruitment is parameterized as

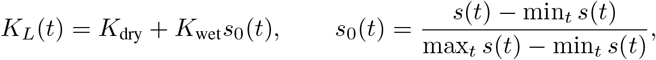

where *s*(*t*) is the normalized monthly seasonality profile and *K*_dry_ is a fixed dry-season larval-recruitment floor. This parametrization keeps the dry-season floor fixed across target EIR levels and changes transmission intensity through the wet-season amplitude. In matched-EIR analyses, *K*_wet_ is calibrated for each posterior/vector draw so the underlying transmission potential before routine case management reproduces the target annual EIR (§3.4.5). In the routine-care decision projections below, this matched vector forcing is then held fixed while background case management is enabled.

### 3.2 Calibration data and likelihood

#### Sites and data

We calibrate jointly to eight site-level data streams spanning a broad transmission range. The five cross-sectional calibration sites are **Namawala 1991** (Tanzania, EIR ≈ 329), **Dielmo 1990** (Senegal, EIR ≈ 200), **Chonyi 1999** (Kenya, EIR ≈ 50), **Ndiop 1993** (Senegal, EIR ≈ 20), and **Ngerenya 1999** (Kenya, EIR ≈ 13). These data provide age-stratified diagnostic-positive prevalence and clinical-incidence summaries, with reported sample sizes and person-time where available; missing denominators are imputed at *n* = 50 per bin. The joint calibration also adds three pre-intervention Garki Project villages (Matsari, Rafin Marke, Sugungum, Nigeria, 1970–1972), using repeated age-stratified prevalence and infectiousness targets after the observation-process correction described in the supplementary Garki calibration appendix.

#### Observation models

We observe two outcomes per site and age bin: diagnostic positivity *µ*_*s,a*_ and clinical incidence 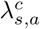 Both are computed as risk-weighted averages over the compartmental state. Diagnostic positivity uses a logistic detection model that increases with MOI and decreases with immunity, age, and maternal protection. Clinical incidence uses an analogous logistic model for the raw clinical probability, then applies a refractory-period correction so repeated clinical days within a single infectious episode are not counted as independent episodes. The calibration likelihood treats clinical incidence as an event output rather than fitting a care-seeking model; routine treatment is not part of posterior calibration. In the decision case study below, routine case management is added as a fixed scenario layer, not as an optimized intervention: symptom-triggered clearance is enabled at 60% for under-fives, 45% for ages 5–15, and 30% for ages 15 and older. In both observation models, uninfected cells (*k* = 0) contribute zero event probability. Exact observation equations, aggregation over biting-risk strata, and parameter definitions are given in the supplementary model-parameter appendix.

The calibrated posterior uses a 38-dimensional parameter vector: 30 shared natural-history and observation scalars plus 8 site-specific EIR adjustments. The shared scalars include immunity acquisition and waning, infection duration, pre-erythrocytic protection, RDT detection, clinical incidence, and the age-, immunity-, and MOI-resolved human-to-mosquito infectiousness kernel. The 8 EIR adjustments comprise five log-scale corrections for the non-Garki calibration sites and three Garki annual-EIR scalars (supplementary Garki calibration appendix). Bounds and priors for all calibrated parameters, plus the fixed structural kernels and prior-construction provenance used by the observation models, are catalogued in the supplementary model-parameter appendix.

#### Likelihood

The calibration likelihood is denominator-aware. Cross-sectional diagnostic positives are modeled with beta-binomial likelihoods, clinical counts with negative-binomial likelihoods over person-time, and Garki infectiousness targets with beta likelihoods centered on reconstructed human-to-vector infectiousness. Full likelihood equations, over-dispersion constants, denominator handling, and the Garki infectiousness likelihood are given in the supplementary NUTS diagnostics appendix.

### 3.3 Intervention and cost model

We implement four interventions with continuous program-level coverage variables ***θ***_int_ = (cov_ITN_, cov_SMC_, cov_IRS_, cov_Vacc_) ∈ [0, 1]^4^. The mechanistic intervention functions are defined over the full unit interval, while the decision optimization below caps coverage at 90% (§3.4.3). Coverage scales the relevant eligible population or program target for each intervention: households or persons for ITNs, eligible children for SMC and vaccination, and resident exposure covered by sprayed structures for IRS. This section focuses on how each intervention modifies the dynamics; Table 1 maps the intervention parameters used in the decision analysis, while full prior definitions, empirical anchors, and prior-predictive consistency figures are given in the supplementary intervention appendices.

**Table 1:** Intervention parameters propagated in the decision analysis. Numeric prior ranges, empirical anchors, and prior-predictive consistency figures are reported in the supplementary intervention appendices.

| Intervention | Sampled efficacy / durability parameters | Fixed programme inputs and shared terms | Cost terms |
| --- | --- | --- | --- |
| ITN | Initial blocking; initial killing; usage conditional on retention; blocking and killing decay; median physical retention | Campaign distribution; indoor exposure from shared vector-behaviour draw | $c_{\text{ITN}}, \gamma_{\text{ITN}}$ |
| IRS | Fresh-substrate kill; field attenuation; operational implementation; residual half-life | Annual spraying; fixed decay steepness; indoor-resting exposure derived from shared vector-behaviour draw | $c_{\text{IRS}}, \gamma_{\text{IRS}}$ |
| SMC | Prophylactic half-life; peak per-dose protection; per-cycle curative clearance | Four monthly Jul–Oct cycles; under-five target population | $c_{\text{SMC}}, \gamma_{\text{SMC}}$ |
| Vaccine | Peak protection against new infection; waning half-life | Under-five target population; fixed deployment timing in the decision sweep | $c_{\text{Vacc}}, \gamma_{\text{Vacc}}$ |

### Insecticide-treated nets (ITN)

#### Mechanism and priors

Insecticide-treated nets reduce successful indoor biting through blocking and increase mosquito mortality through kill-on-feed. Following the mechanism frameworks of Killeen et al. [2007] and Sherrard-Smith et al. [2022], blocking and killing decay after distribution, while net retention follows a Weibull survival curve with median retention *T*_50_ [Bertozzi-Villa et al., 2021]. Effective net exposure is the product of campaign coverage, use conditional on retention, and physical retention. The indoor-night biting fraction belongs to the shared vector-behavioural layer and is therefore drawn once and reused for the ITN and IRS mechanisms. Full prior ranges and empirical anchors are reported in the supplementary ITN appendix.

### Indoor residual spraying (IRS)

#### Mechanism and priors

IRS kills mosquitoes that feed indoors and then remain indoors long enough to rest on treated surfaces. The fresh-spray kill probability combines laboratory cone-bioassay efficacy, field attenuation, and operational implementation, then decays within each annual spray cycle according to a residual half-life. The indoor-resting exposure term is derived from the same indoor-night biting draw used by the ITN module and an early-exit fraction, enforcing indoor resting to be no greater than indoor feeding. Substrate-specific anchors and ranges are reported in the supplementary IRS appendix.

#### 3.3.1 Combined ITN + IRS deployment

ITNs and IRS act on overlapping subsets of mosquitoes: both act on mosquitoes that attempt to feed indoors, while IRS additionally requires the mosquito to remain indoors long enough to rest on a sprayed surface. Naively summing the mortality increments from the two interventions would double-count this overlap, because a mosquito killed on the net never reaches a sprayed wall. We therefore use a **sequential** (ITN → IRS) composition in the spirit of Le Menach et al. [2007] and the Imperial framework of Griffin et al. [2010]: the IRS mortality contribution is discounted by the fraction of indoor-feeding mosquitoes that survive the net encounter. IRS does not deter biting in the model, so the biting-reduction channel is unchanged from the ITN-only expression. The composition recovers the ITN-only and IRS-only limits when the other coverage is zero; algebraic checks and the magnitude of the naive-additivity discount are given in the supplementary IRS appendix.

### Seasonal malaria chemoprevention (SMC)

#### Mechanism and priors

SMC is modelled as four monthly SP+AQ cycles delivered during the peak-transmission season to children under 5 y, the age-grid approximation to the usual 3–59 mo target. Each cycle has a curative component that clears a fraction of existing parasitemia in eligible children and a prophylactic component that reduces new infections and wanes between cycles. The Jul–Oct schedule and under-five target are fixed programme inputs. Prior definitions and prior-predictive comparisons with trial and programme-effectiveness evidence are reported in the supplementary SMC appendix.

### R21/RTS,S vaccine

#### Mechanism and priors

Vaccination is modelled as waning prophylactic protection against new infection in children under 5 y, with no curative clearance arm. The decision-analysis prior is a generic modern pre-erythrocytic vaccine prior spanning RTS,S/AS01 and R21/Matrix-M, rather than a product-specific vaccine model; deployment timing and the under-five target are fixed programme inputs. The R21-specific prior-predictive check uses product-specific protection priors and is reported in the supplementary vaccine appendix.

#### Cost model

Decision budgets are expressed as US dollars per resident over the 3-year optimization horizon. We use a convex resident-equivalent cost model,

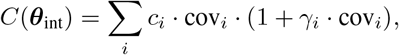

where *c*_*i*_ is the base resident-equivalent cost coefficient after any target-population scaling, so full coverage costs *c*_*i*_(1 + *γ*_*i*_). The coefficient *γ*_*i*_ is an operational-convexity planning prior capturing increasing delivery overhead at high coverage. Base-cost priors are lognormal and overhead priors are normal, anchored to published delivery-cost ranges; intervention-specific medians, uncertainty ranges, and scaling conventions are given in the supplementary cost-prior appendix.

Intervention efficacy, durability, exposure, and cost priors are literature-anchored planning assumptions for the decision analysis. They are not updated by the calibration likelihood. Effect sizes obtained by propagating these priors through the forward model are compared with published RCT and programme-effectiveness estimates in the supplementary intervention appendices.

### 3.4 Inference and decision analysis

#### 3.4.1 Differentiable implementation

Delenda is implemented end-to-end in JAX [Bradbury et al., 2018], with the forward map a single `jax.jit`-compiled function from (*θ*, ***θ***_int_, *θ*_vec_) to all observables. The dominant cost is the inner ODE integration, so we use a custom vector–Jacobian product (VJP) first-order adjoint for the solver [Chen et al., 2018]. The result behaves as a standard JAX primitive: it is vmappable over sites and posterior draws, reverse-mode differentiable for likelihood gradients and intervention-mix optimization, and JIT-compilable to a single XLA graph. These gradients support both posterior estimation and continuous intervention optimization, while the completed NUTS run remains the posterior of record. Implementation details and the memory tradeoff are given in the supplementary differentiable-implementation appendix.

#### 3.4.2 Posterior estimation

We use stochastic variational inference (SVI) as computational scaffolding rather than as the final uncertainty estimate. A full-rank Gaussian guide completes in roughly one hour and supplies rapid development checks, initialization, and mass-matrix geometry for the Hamiltonian Monte Carlo run. The posterior of record is then sampled with the No-U-Turn Sampler (NUTS; Hoffman and Gelman 2014), targeting the bounded-parameter posterior itself. The production NUTS run took roughly two weeks on a single GPU. The supplementary NUTS diagnostics appendix reports the bounded-parameter transform, sampler configuration, diagnostics, and comparison with the SVI approximation.

#### 3.4.3 Bayesian intervention-mix optimization

For decision-support, we optimize continuous intervention coverage ***θ***_int_ ∈ [0, 0.9]^4^ for ITN, SMC, IRS, and vaccine coverage. The 90% upper bound avoids treating perfect coverage as an operationally attainable state. For each target EIR, budget, uncertainty stage, and objective, we solve

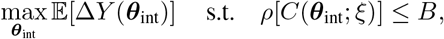

where Δ*Y* is either under-five or all-age clinical cases averted over the 3-year horizon, *B* ∈ {1, 2, 3, 5, 7, 10, 15, 20, 25} dollars per resident, and *ρ* is the budget rule. We use three budget rules: expected cost (*ρ*[*C*] = *E* [*C*]), the 90th percentile cost (*q*_0.90_[*C*]), and CVaR90 [Rockafellar and Uryasev, 2000], the mean cost in the most expensive 10% of cost-prior draws. Automatic differentiation supplies gradients of the disease objective through the JAX forward model, while the sampled cost functions are analytic in coverage. This lets the optimizer search the continuous four-intervention coverage space directly under each budget rule.

Combining the NUTS posterior with literature-anchored intervention, vector-biology, and cost priors yields the joint uncertainty distribution used to propagate intervention uncertainty alongside model uncertainty. Positive quantities such as costs, half-lives, and decay timescales use lognormal priors; probabilities and coverage modifiers use truncated-normal priors on [0, 1]. Each decision cell uses draw-level paired simulations: the baseline scenario and the intervention scenario share the same model, vector, and intervention draw, so cases averted are computed as a paired contrast. In the application below, the baseline includes fixed routine case management while the four optimized prevention interventions are set to zero.

#### 3.4.4 Vector biology priors

The vector-behavioural layer comprises seven parameters that are not calibrated to data; each receives a literature-anchored prior with central values consistent with *An. gambiae sensu lato* in West African endemic settings. These priors cover host seeking, anthropophily, mosquito mortality, extrinsic incubation period, vector-to-human transmission, indoor night biting, and early exit after feeding. The supplement gives the full prior intervals and prior-predictive diagnostics.

Two of these parameters describe where biting and resting happen relative to indoor surfaces. We model these in the vector layer so that ITNs and IRS share the same draw of the indoor-feeding fraction INDOOR_*n*_, while the indoor-resting fraction used by IRS is derived as INDOOR_*r*_ = INDOOR_*n*_(1 − *ρ*_exit_). This construction enforces the biological hierarchy INDOOR_*r*_ ≤ INDOOR_*n*_. At the joint median, INDOOR_*r*_ = 0.60 · 0.90 = 0.54.

#### 3.4.5 Conditioning on observed EIR

Vector-behavioural parameters and seasonal larval recruitment jointly determine the EIR faced by the human population. For decision projections, I therefore condition directly on the target annual EIR: for each draw of the calibrated model parameters and vector priors, the wet-season larval-recruitment amplitude *K*_wet_ is chosen so that the simulated baseline reproduces the target EIR.

This conditioning matters for uncertainty attribution. Without re-solving *K*_wet_, vector-ecology draws would also move the realized baseline EIR, mixing vector biology with transmission-intensity drift. Matched-EIR conditioning removes that drift, so vector-biological uncertainty is assessed conditional on the observed site-level EIR rather than confounded with it; the purpose is to isolate vector uncertainty, not to remove it. The supplement gives the seasonal *K*_*L*_ parameterization, numerical solve, status codes, and prior-predictive feasibility checks used to implement this conditioning.

#### 3.4.6 Illustrative seasonal setting and routine treatment

The decision case study uses the monthly EIR seasonality profile from Dapelogo, Burkina Faso, as an illustrative highly seasonal forcing profile. The profile is normalized to define the monthly shape *s*_0_(*t*) in *K*_*L*_(*t*), while the absolute transmission intensity is set by the matched-EIR procedure above. The Dapelogo profile is therefore used to represent a plausible dry-season/wet-season contrast, not to make a deployment recommendation for Dapelogo itself.

The decision simulations also include fixed routine case management. After *K*_wet_ is matched to the underlying transmission potential before case management, the vector environment is held fixed and routine treatment is enabled during the burn-in and intervention horizon. This fixed-vector convention keeps the EIR label interpretable as underlying transmission potential before case management. Treatment seeking is implemented as symptom-triggered parasite clearance: for age *a*, immunity *u*, and MOI *k*, the clearance rate is *h*_clin_(*a, u, k*)*f*_treat_(*a*), where *h*_clin_ is the calibrated clinical hazard and *f*_treat_(*a*) is a fixed age-specific care-seeking probability. Treated infected individuals move to MOI *k* = 0 without losing latent immunity. The routine-treatment scenario uses *f*_treat_ = 0.60 for ages < 5 years, 0.45 for ages 5–15 years, and 0.30 for ages ≥15 years. ITNs, SMC, IRS, and vaccine coverage are then optimized as added prevention interventions on top of this routine-care baseline.

## 4 Application: Highly seasonal cost-constrained intervention optimization

We apply the framework to an illustrative, highly seasonal transmission scenario parameterized around Dapelogo, Burkina Faso; the routine-care baseline and seasonal profile are shown in the supplementary Dapelogo-baseline appendix and defined in §3.4.6. **This is a methodological case study, not a deployment-ready recommendation**: the optimized portfolios show how intervention priorities shift under different transmission, cost, age-weighting, and uncertainty assumptions, not what any district should deploy. Intervention efficacy and cost parameters are planning-scale literature priors rather than application-specific programme inputs, and vector biology is not site-elicited; the local inputs an operational analysis would require are enumerated in the Caveats (§4.6).

The case study reuses the calibrated model rather than re-fitting it. We draw the shared natural-history, immunity, detection, clinical, and infectiousness parameters from the eight-site posterior; the per-site EIR terms are specific to the calibration sites and are dropped, replaced by a swept target EIR that places the same biology across a range of transmission intensities. Calibration drives each site by a fixed EIR input with a calibrated per-site log-scale correction; the decision analysis instead runs the closed-loop human–vector system and re-solves the seasonal larval amplitude *K*_wet_ so that each posterior/vector draw reproduces the target EIR under the Dapelogo seasonal profile (§3.4.5). Calibration thus constrains the biology; the decision sweep exercises that biology in a new transmission, budget, and objective context.

Unless otherwise stated, the decision figures in this case study use the same design: the seasonal vector environment is first matched to the target EIR before routine case management is applied, then held fixed while routine case management is enabled during a 60-year burn-in and 3-year intervention horizon. Target EIR values are {1, 3, 10, 30, 100} infectious bites per person-year, and resident-equivalent budget values on the grid {1, 2, 3, 5, 7, 10, 15, 20, 25} US dollars per resident. The near-elimination end of this sweep (EIR 1–3) lies below the lowest-transmission calibration site, so those cells are extrapolations below the calibrated range. Stochastic uncertainty rows use 100 paired draw-level simulations per EIR–budget cell, and optimized coverages are continuous but capped at 90%. Uncertainty is added in five cumulative stages—a median-input baseline, then posterior, vector-biology, intervention-efficacy, and cost uncertainty in turn (Table 2).

**Table 2:** Uncertainty sources propagated in the decision-analysis workflow. The calibrated posterior controls natural-history and site-EIR uncertainty; the other sources are literature-anchored planning priors.

| Source | Random quantities | Where it enters | Role in analysis |
| --- | --- | --- | --- |
| Calibrated posterior | 38 calibrated human-model and site-EIR parameters | Infection, immunity, detection, clinical incidence, infectiousness, and site calibration | Biological uncertainty from the NUTS posterior |
| Vector biology | Biting, anthropophily, mortality, EIP, V2H, indoor biting, early exit | Vector equilibrium; matched-EIR conditioning; ITN/IRS mechanisms | Vector-behaviour uncertainty conditional on observed EIR |
| Intervention efficacy | ITN, IRS, SMC, and vaccine effect and waning parameters | Intervention schedules; modified force of infection and vector mortality | Product and delivery-effectiveness uncertainty |
| Cost | Resident-equivalent unit costs; high-coverage overhead | Budget constraint | Median, expected-cost, and cost-risk budget analyses |

The application proceeds in five steps. We first show the calibrated posterior predictive fit, then report the computational scale of the decision workflow. We then optimize expected-cost portfolios, examine whether uncertainty changes portfolios or primarily changes impact distributions, and finally replace the expected-cost constraint with a tail-risk constraint to quantify the impact–budget-risk tradeoff.

### 4.1 Calibration results

Figure 2 shows the eight-site NUTS posterior predictive from the completed production NUTS run. The calibration jointly fits five cross-sectional sites together with three additional pre-intervention villages from the Garki Project (Matsari, Rafin Marke, and Sugungum, Nigeria, 1970–1972). The supplementary Garki calibration appendix describes the Garki design and the microscopy-sensitivity correction used to place Garki prevalence on a scale comparable to the other calibration sites. The calibrated parameter vector has 38 entries: 30 shared dynamics, detection, clinical, recovery, protection, and infectiousness-kernel parameters, plus site-specific EIR adjustments for the five cross-sectional sites and three Garki EIR scalars *κ*_*s*_ centered on the Garki Monograph values.

**Figure 1:**
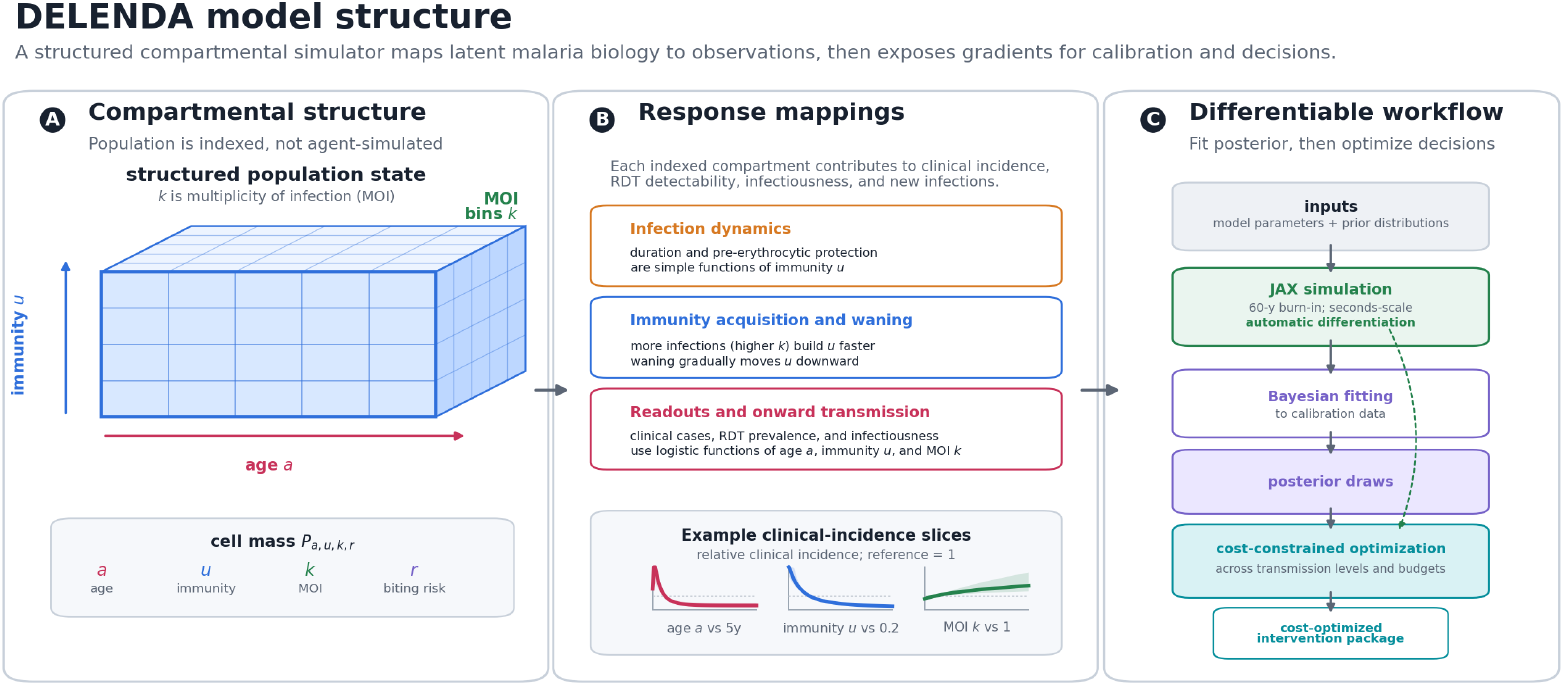
Delenda model structure and differentiable workflow. The model tracks population mass over age, latent immunity, multiplicity of infection (MOI), and biting-risk strata rather than simulating individual histories. Response mappings convert each indexed compartment into infection dynamics, immunity acquisition and waning, clinical incidence, RDT prevalence, and infectiousness to mosquitoes. The same JAX implementation is then used first for Bayesian fitting to calibration data and subsequently for cost-constrained intervention optimization across transmission levels and budgets.

**Figure 2:**
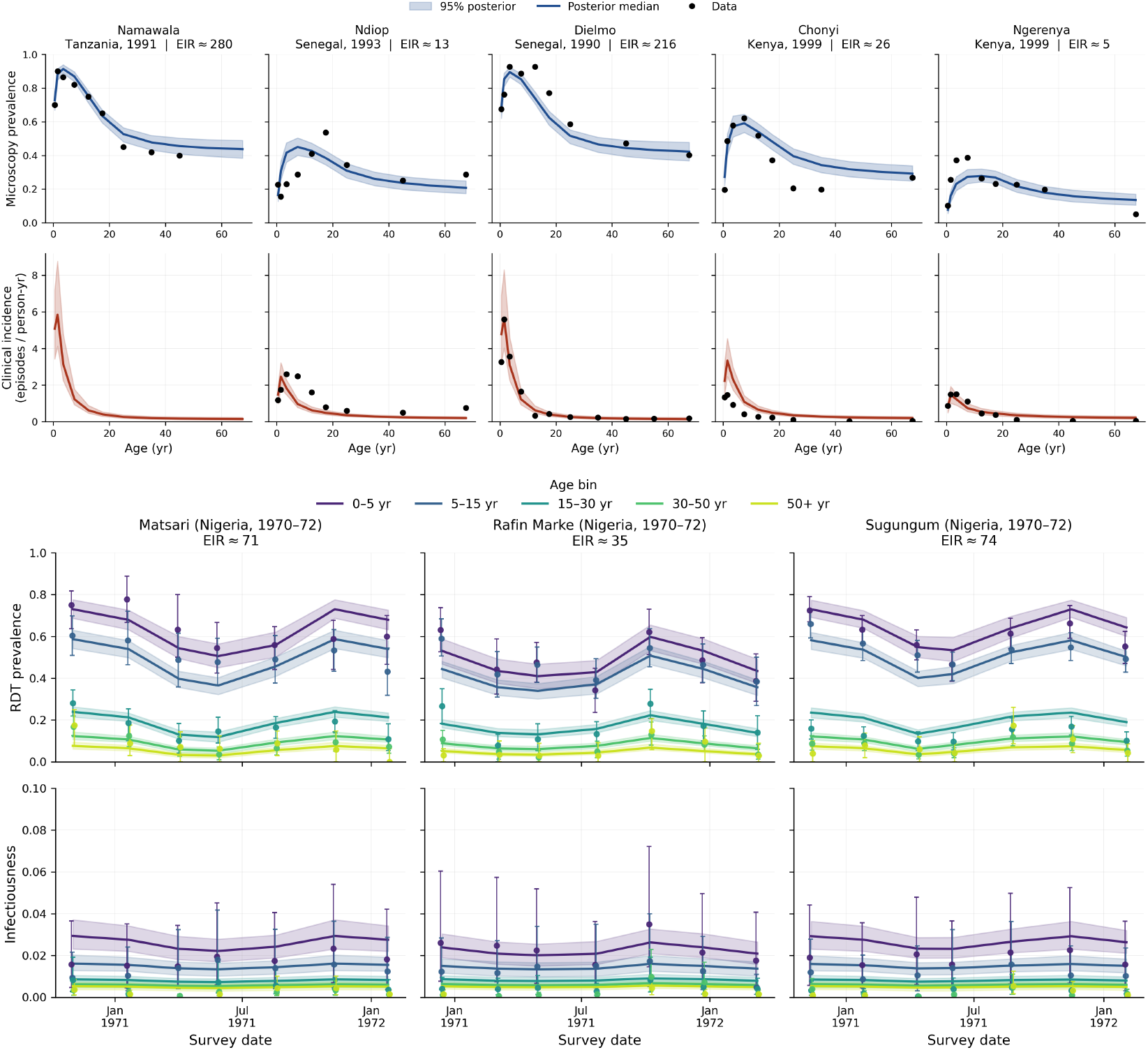
Eight-site NUTS posterior predictive calibration. Top panel: five cross-sectional calibration sites, with microscopy/RDT prevalence by age above clinical incidence per person-year. Bottom panel: three Garki Project villages (Nigeria, 1970–1972), with standard-microscopy-equivalent prevalence above expected human-to-mosquito infectiousness. Bands are 95% credible intervals from 200 draws of the completed NUTS posterior; lines are posterior medians and points are calibration targets with uncertainty intervals where available.

The posterior predictive bands track age-prevalence and age-incidence across the cross-sectional EIR range while also reproducing the seasonally resolved Garki prevalence and infectiousness targets. As shown in the supplementary NUTS diagnostics appendix, the NUTS posterior and the SVI-preconditioned guide lead to the same predictive regime, with NUTS yielding appropriately wider non-variational uncertainty bands.

The posterior narrows several central scale and readout parameters while leaving some shape, maturation, and site-EIR directions weakly informed; the supplementary model-parameter appendix reports the parameter-level prior-vs-posterior intervals and mechanism curves.

### 4.2 Computational performance

Computational efficiency is central to Delenda‘s contribution, and it follows from a compact compartmental structure made differentiable in JAX. The same compiled forward map is reused for 60-year burn-ins, vectorized posterior-predictive checks, reverse-mode calibration gradients, and gradient-based intervention optimization on one GPU. Because that map is differentiable, continuous coverage is optimized directly rather than by coverage-grid enumeration, while posterior, vector, intervention, and cost uncertainty are propagated by vectorized draw-level simulation.

Table 3 reports empirical timings from the calibration and decision runs. The expensive decision step is preparing draw-specific baselines: each posterior/vector draw is matched to a target EIR after a 60-year burn-in. The workflow pays this cost once for each target EIR and model/vector uncertainty stage, then amortizes it by caching the matched equilibrium states. The cached states are reused across budgets, objectives, intervention-prior draws, and cost-risk rules. After this conditioning step, each decision cell reduces to a four-variable continuous constrained optimization over ITN, SMC, IRS, and vaccine coverage. JAX supplies gradients of the health objective, so the optimizer can search the continuous coverage space directly rather than evaluating a combinatorial grid of candidate portfolios.

**Table 3:** Computational scale of posterior fitting and decision analysis. Timings are empirical wall-clock summaries from the single-GPU workflow. Decision runs use 100 posterior/vector draws, 60-year burn-in, five EIR levels, nine budgets, five uncertainty stages, and two objective functions. Per-cell optimization times exclude the separately reported EIR-matching step.

| Operation | Scale | Empirical timing |
| --- | --- | --- |
| SVI preconditioner | Full-rank Gaussian guide for the 38-parameter posterior | Approximately 1 h |
| Production NUTS posterior | 4 chains, 2160 post-warmup draws total | Approximately 2 weeks |
| First-pass EIR matching | 100 draws for one target EIR and one model/vector stage | Median 19.7 min per EIR–stage fit |
| Cached EIR reuse | Same matched draw set reused across intervention and cost-prior stages | Median 0.2 s per EIR–stage cache hit |
| Expected-cost optimization | 450 continuous optimization cells | 0 failures; median 31.6 s/cell; 90th percentile 78.1 s/cell |
| 90th-percentile-cost optimization | 450 continuous optimization cells | 0 failures; median 34.9 s/cell; 90th percentile 108.9 s/cell |
| CVaR90-budget optimization | 450 continuous optimization cells | 0 failures; median 29.6 s/cell; 90th percentile 82.0 s/cell |
| Full decision-figure refresh | Routine-treatment expected-cost, cost-risk, baseline, single-intervention, and efficacy-robustness outputs | Approximately 1 day |

### 4.3 Expected-cost intervention portfolios

We first optimize under the expected-cost budget rule, *E*[*C*(*θ*)] ≤ *B*. Figure 3 summarizes the full-uncertainty expected-cost optima across the shared EIR and budget grid for both under-five and all-age cases averted.

**Figure 3:**
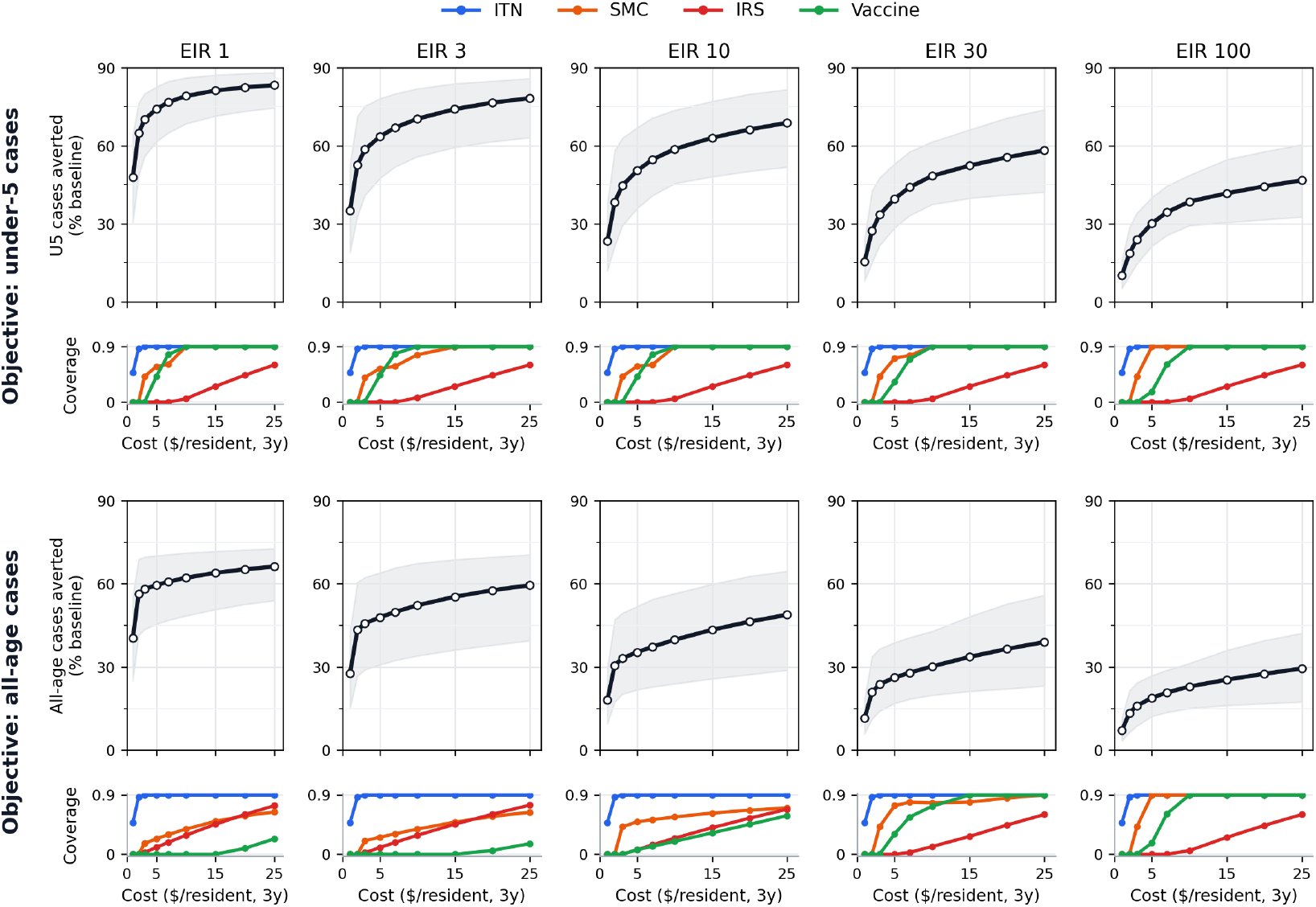
Expected-cost optimized portfolio frontier. Each column fixes the matched annual EIR and traces the optimized portfolio as the 3-year resident-equivalent budget increases. Upper panels show the percentage of baseline clinical cases averted by the optimized portfolio; gray ribbons show central 95% intervals across paired posterior, vector, and intervention-efficacy outcome draws. Lower panels show the corresponding optimized continuous coverage for ITNs, SMC, IRS, and pediatric vaccination, with maximum coverage capped at 90%. The top row maximizes under-five cases averted; the bottom row maximizes all-age cases averted.

The first-order intervention ranking is stable. ITNs enter first and reach the 90% coverage cap by budgets of roughly $3–$5 across the EIR sweep. SMC then saturates rapidly for the under-five objective, while vaccine coverage enters at moderate budgets and IRS appears later as an expensive add-on once cheaper child-targeted and net-based protection is near saturation. The all-age objective shifts allocation in the expected direction: vaccination is delayed or reduced where its child-targeted protection is diluted by the under-five population fraction, and IRS receives more weight because it protects all ages through vector mortality. These objective-driven differences are larger than the coverage changes induced by uncertainty, which we quantify next (§4.4). This ranking stability partly reflects wide separation in marginal cost-effectiveness and age-targeting across the intervention classes. The supplementary single-intervention cost-effect diagnostic provides marginal intuition for the optimized multi-intervention frontier in Figure 3.

### 4.4 Uncertainty changes impact more than decisions

The robust-ranking result does not mean uncertainty is unimportant. Uncertainty enters the decision analysis mainly through the *distribution of impact*, not through frequent changes in the optimized coverage mix. Relative to the median-input portfolio, adding all biological and intervention uncertainty shifts optimized coverage only modestly. Here L1 coverage shift means the sum of absolute differences across the four optimized coverages; for example, a value of 0.05 corresponds to five percentage points of total coverage moved across the portfolio. The mean L1 coverage shift is 0.036 for the under-five objective and 0.049 for the all-age objective in the expected-cost full-uncertainty run. The largest single-cell L1 shift is 0.085 for under-five and 0.236 for all-age, both well below a wholesale change in intervention ranking.

Figure 4 shows this separation. Optimized allocation curves largely overlap as posterior, vector, efficacy, and cost uncertainty are added, while the full-uncertainty impact intervals remain wide. In the full-uncertainty run, the median draw-level standard deviation across EIR–budget cells is 6.5 percentage points for under-five cases averted and 7.3 percentage points for all-age cases averted. Thus the posterior-aware workflow does not primarily say “choose a different package.” It quantifies how wide the impact distribution is around the package that remains optimal.

**Figure 4:**
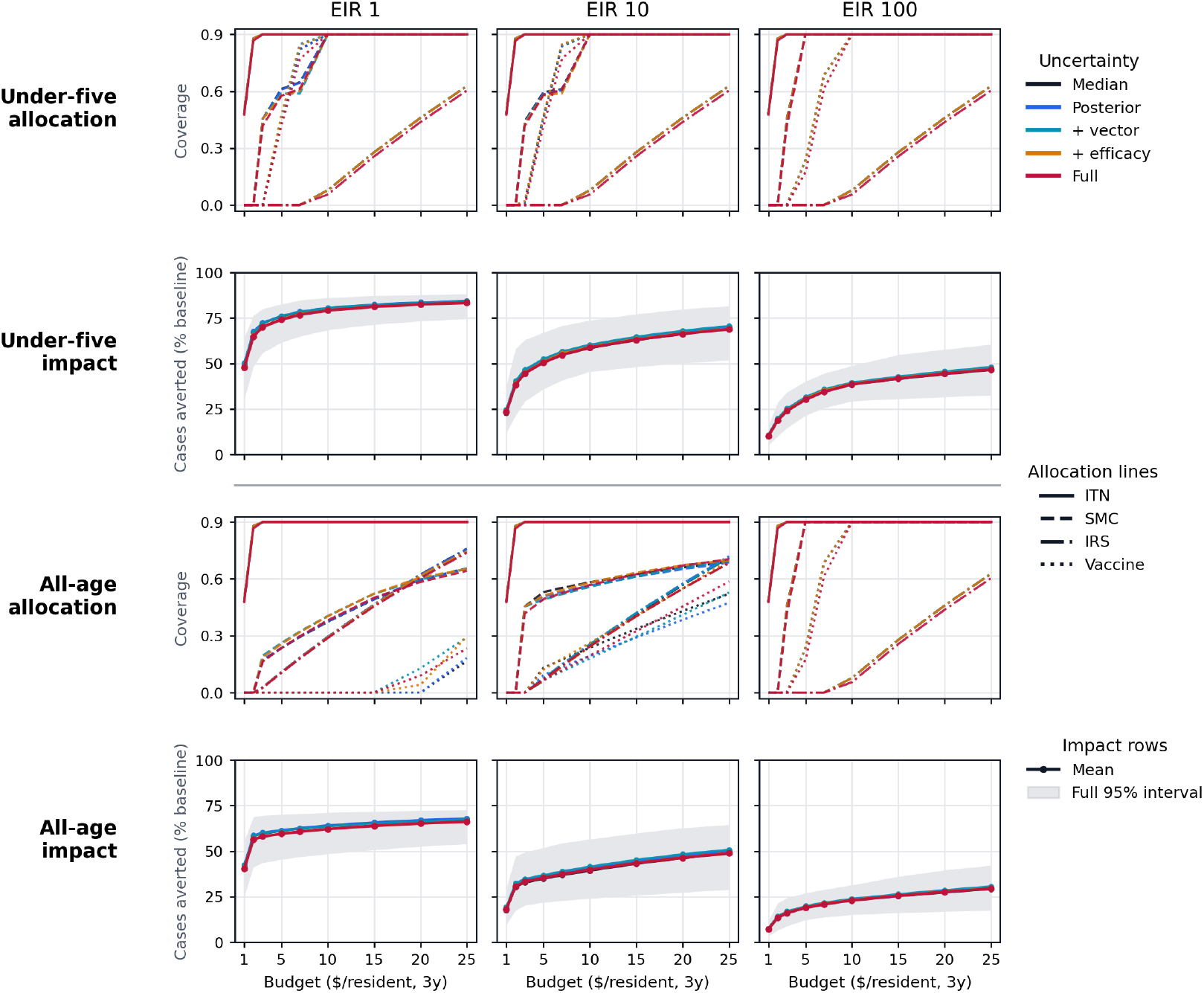
Uncertainty changes impact more than optimized allocation. Columns show representative low, intermediate, and high matched annual EIR values. Allocation rows plot the expected-cost optimized coverage as budget increases; colors add uncertainty sources sequentially and line styles denote intervention classes. Impact rows plot mean cases averted as a percentage of the routine-care baseline for the corresponding optimized portfolio; the grey band is the full-uncertainty central 95% interval.

### 4.5 Decisions under cost uncertainty

Expected-cost optimization treats the average sampled cost as the budget constraint. This is useful for estimating expected impact, but it leaves substantial budget-overrun risk: in the full-uncertainty expected-cost run, the probability that realized cost exceeds the nominal budget ranges from 42% to 52% across optimized cells. We quantify the constraint-side effect of cost uncertainty by solving the same continuous optimization problem under two tail-risk budget rules. The q90 rule constrains the empirical 90th percentile of sampled costs, while the CVaR90 rule constrains the mean cost in the most expensive 10% of cost-prior draws. In finite samples, q90 corresponds to allowing roughly one in ten cost draws to exceed the nominal budget; CVaR90 is stricter because it controls the average severity of the upper tail.

Table 4 shows that risk-aware budget rules retain the same qualitative ranking but trim coverage to buy budget protection. q90 optimization reduces the average overrun probability to approximately 10%, with an 8.2–8.4% mean impact penalty relative to expected-cost optimization. CVaR90 is more conservative, reducing average overrun probability to 4.4–4.6% with a 10.9–11.2% mean impact penalty. At a $7 budget, for example, under-five optimization at EIR 100 selects ITN=0.90, SMC=0.90, IRS=0, and vaccine=0.25 under CVaR90 rather than vaccine=0.61 under the expected-cost rule. The risk-aware analysis is therefore best read as an operating margin: how much expected impact a program is willing to sacrifice to reduce the probability and severity of budget overrun.

**Table 4:** Budget-risk rules trade expected impact for lower overrun probability. Summary across the 45 full-uncertainty EIR–budget cells for each objective. Impact penalty is the mean relative reduction in optimized cases averted compared with the expected-cost optimum for the same EIR, budget, and objective. L1 shift is the summed absolute coverage change across ITN, SMC, IRS, and vaccine coverage. Dashes mark the expected-cost comparator.

| Budget rule | Objective | Budget-overrun probability | Impact penalty | L1 coverage shift vs expected-cost policy |
| --- | --- | --- | --- | --- |
| Expected cost | Under-five | 45.7% | – | – |
| q90 | Under-five | 10.0% | 8.2% | 0.21 |
| CVaR90 | Under-five | 4.6% | 10.9% | 0.27 |
| Expected cost | All-age | 46.8% | – | – |
| q90 | All-age | 9.8% | 8.4% | 0.27 |
| CVaR90 | All-age | 4.4% | 11.2% | 0.27 |

### 4.6 Caveats

Several limitations should shape how the proof-of-concept results are read.

1. The joint posterior includes Garki-derived infectiousness targets, but those targets still depend on a density-to-infectiousness observation model and sparse historical gametocyte data. Direct calibration to modern paired density/DMFA datasets remains a natural extension.
2. Cost and most efficacy parameters are planning-scale literature priors rather than application-specific programme inputs. Operational use would require local procurement, delivery, wastage, product availability, resistance, efficacy elicitation, current coverage, implementation constraints, and programme objectives.
3. The intervention-efficacy priors are not updated by the calibration likelihood. To test whether this matters for the decisions, the supplementary efficacy-robustness appendix re-optimizes portfolios after shifting efficacy priors to adverse literature-supported values. Across 198 EIR–budget–objective cells, the maximum deployment regret is 2.77% and no cell exceeds the 5% materiality threshold, so the tested misspecifications change projected impact more than they change the actionable portfolio.
4. The robust-ranking result is empirical, not guaranteed. Settings with different intervention prices, current coverage, product availability, vector composition, or stronger nonlinear interactions could produce more frequent portfolio changes under uncertainty.
5. Vector dynamics are modeled as a single *An. gambiae s*.*l*.-like species. Settings with multiple vector species that have substantially different biting or resting behavior could shift intervention rankings, particularly the IRS-vs-ITN tradeoff. Under the current cost and efficacy priors, ITNs remain difficult to displace because they are substantially more cost-effective for indoor biting transmission; that ranking should still be rechecked with local vector species composition, costs, and resistance. Adding a second species block is a straightforward parametric extension and is not pursued here.

## 5 Discussion

### Why differentiability changes the workflow

The main architectural claim is not that compartmental models should replace individual-based malaria simulators. Agent-based models remain the natural choice when household structure, individual histories, spatial mobility, or implementation processes are themselves the object of study. Our claim is narrower: many intervention-prioritization questions in endemic settings are population-level allocation problems. For those, a structured compartmental model can preserve enough malaria mechanism while making posterior inference, uncertainty propagation, and optimization routine rather than exceptional. The key computational move is not merely making the simulator smaller; it is making the simulator differentiable. In non-gradient individual-based workflows, computational cost often forces analyses to expose only a small subset of parameters and to rely on scenario sweeps or derivative-free search. In Delenda, the same JAX forward model supports likelihood-based calibration, posterior sampling, matched-EIR conditioning, and continuous constrained optimization. The custom adjoint is an implementation detail that makes this differentiable workflow practical at the current state size by keeping memory use under control during long burn-ins and intervention projections. We expect this pattern—mechanistic compartmental model + automatic differentiation + posterior-aware constrained optimization—to generalize beyond malaria to many other infectious-disease and chronic-disease control problems.

### What posterior-aware optimization buys

Carrying posterior and prior uncertainty into the optimizer changes the allocation problem in two separable ways. First, it turns projected impact into a distribution rather than a point estimate. Second, it tests whether that uncertainty lies along directions that actually alter the optimal portfolio. In the highly seasonal decision grid it does the former far more than the latter: posterior, vector, and efficacy uncertainty widen the impact distribution much more than they change intervention rankings—a useful result, not a failure of the analysis. The portfolio is stable under the modeled biological uncertainties while the magnitude of impact remains uncertain.

### Where uncertainty matters

Three findings from the case study deserve emphasis. First, the optimized portfolios are more robust than their projected impact: posterior, vector, and efficacy uncertainty change the coverage mix only modestly—under the modeled cost and efficacy priors the optima are ITN-heavy across the grid—while widening the distribution of cases averted (§4.4). Second, the under-five-versus-all-age objective is a larger qualitative lever than that uncertainty: child-targeted SMC and vaccination enter earlier under the under-five objective, and broader population protection is favored under all-age optimization. Third, cost uncertainty acts mainly on the constraint: expected-cost optima carry a material budget-overrun probability that tail-risk rules buy down at a modest impact cost (§4.5). None of these required model elaboration; they followed from running the same calibrated model under different objectives and budget-risk rules.

### How to read the optimized portfolios

The claim is not that the displayed portfolios are the right choices for Dapelogo, but that a posterior-aware compartmental model can turn a high-level SNT question into a transparent constrained optimization and report both the portfolio and its carried uncertainty. Robustness here means robustness over the calibrated posterior and the specified vector, efficacy, and cost-prior layers; it is not a claim of invariance to all structural-model alternatives or local implementation constraints.

### Future work

Several extensions would move this framework toward an operational SNT workflow. First, intervention realism can be increased without changing the core decision formulation. For example, pyrethroid-only, PBO, and dual-active-ingredient nets can be represented as alternative ITN parameterizations with different kill, durability, and cost priors; the optimizer would then choose both coverage and product. ITN use can also be made age- and season-specific using survey-derived usage profiles, and the vector-prior layer can be made temperature- and site-specific rather than relying on broad ecological priors.

Second, the single-site optimization should be expanded to the multi-site questions that national malaria programs actually face. A panel of matched-EIR sites would produce a stratification table of optimal packages, posterior-weighted impact, and budget requirement. A shared-budget version would then allocate a fixed national or grant-level budget across sites, reusing the same gradient infrastructure but solving a higher-dimensional allocation problem.

Third, the observation model should move closer to routine data. Matched-EIR conditioning is useful for separating vector uncertainty from baseline transmission, but most programs observe clinical incidence more readily than EIR. Calibrating or conditioning on clinical incidence would test how much additional uncertainty enters when the observation is immunity-modulated, care-seeking dependent, and partially saturated. A related biological extension is to benchmark or refit the infectiousness observation model against modern paired density/DMFA datasets.

Finally, Delenda should be benchmarked against established malaria simulators in a full end-to-end SNT workflow, not only on matched forward-simulation scenarios. A useful comparison would ask whether Delenda and an existing SNT model such as EMOD produce similar stratification tables, intervention rankings, and budget-constrained packages when given the same intervention menu and local inputs, while also comparing runtime, calibration burden, exposed parameter dimension, uncertainty propagation, and optimization behavior. Such comparisons would not be needed to justify every SNT use case, but they would help delimit where a differentiable compartmental model is sufficient and where the additional complexity of individual histories, household structure, or spatial implementation processes truly changes the relevant decision. For large-scale deployment across thousands of sites, neural or other statistical surrogates trained on posterior draws could further accelerate the forward solver while preserving the posterior-aware optimization workflow.

## Data Availability

All code, model definitions, analysis scripts, and manuscript-generation files are available at https://github.com/jgsuresh/compartmental-malaria/tree/medrxiv-2026-07-13. Data used for calibration and intervention-prior checks are derived from previously published sources cited in the manuscript and from repository files where redistribution is permitted. No new human-subject data were collected for this study.

## Acknowledgements

I thank Monique Ambrose for feedback on model design and validation.

## Submission Declarations

### Ethics approval and clinical-trial registration

This is a computational modeling study using published aggregate calibration targets, literature-derived priors, and simulated outputs. No new human participant data were collected, no individual-level or identifiable participant data are analyzed, and no prospective intervention is assigned; institutional ethics review and clinical-trial registration were therefore not required.

### Code and data availability

Code, figure-generation scripts, and processed analysis artifacts will be made available through the project GitHub repository https://github.com/jgsuresh/compartmental-malaria. The submitted version is identified by the release tag `medrxiv-2026-07-13`, and the repository will document the commands used to regenerate the manuscript figures and decision-analysis summaries.

### Licensing

Code is released under the MIT License. Manuscript text, figures, documentation, and generated non-code artifacts are released under CC BY 4.0 unless otherwise noted. Source datasets and third-party materials remain subject to their original licenses and access terms.

## Notes

### Competing Interest Statement

The authors have declared no competing interest.

### Funding

- Gates Foundation

