## Supplement for "DELENDA: Differentiable Epidemiology for Latent-state Estimation and Nonlinear Decision Analysis"

*DELENDA is named after the Gates Foundation motto *Malaria delenda est* ("malaria must be destroyed"), adapted from Cato the Elder's *Carthago delenda est*.

Preprint.

#### Supplementary contents

#### Contents

**A Compartmental model parameters** **3**

**B Differentiable implementation details** **12**

**C Matched-EIR implementation details** **12**

**D Illustrative Dapelogo-inspired routine-care baseline** **13**

**E Intervention cost-prior details** **14**

**F Single-intervention cost-effect diagnostics** **15**

**G Intervention effect-size consistency checks** **19**

**H ITN priors and effect-size check** **19**

**I IRS priors and effect-size check** **20**

**J SMC priors and effect-size check** **21**

**K Vaccine priors and effect-size check** **21**

**L Robustness of the optimized portfolios to intervention efficacy misspecification** **22**

**M Garki Project calibration extension** **23**

**N Posterior estimation and NUTS diagnostics** **25**

### A Compartmental model parameters

This appendix catalogues every parameter that enters the compartmental forward model, separating (i) the 38 scalars estimated in the eight-site NUTS posterior (Table 2; comprising 23 shared dynamics/detection/clinical parameters, 7 infectiousness-kernel parameters calibrated in the eight-site fit, and 8 per-site EIR adjustments) and (ii) the fixed structural constants, response-function kernels, and vector priors that are not estimated in the calibration but are needed to reproduce the model (Table 3). Intervention parameters are covered in the main-text intervention section; only the natural-history parameters appear here. The main-text structure schematic is intentionally conceptual; the tables below give the exact equations, fitted parameters, and fixed constants needed to reproduce the compartmental model.

##### Calibrated parameters (38)

All 38 calibrated parameters are represented on bounded natural scales with truncated-Gaussian priors and sampled in unconstrained logit coordinates by the completed NUTS run (Appendix N). The intervals below report the NUTS posterior; prior-vs-posterior mechanism figures use 1000 draws from the same canonical NUTS sample.

The 38 parameters comprise: 23 shared dynamics, detection, and clinical scalars, the 7-parameter age-, immunity-, and MOI-resolved infectiousness kernel calibrated in the eight-site model, and 8 per-site EIR-adjustment scalars (Table 2).

A CI whose endpoints sit at (or very near) the parameter's bounds indicates that the eight sites do not informatively constrain that parameter under the current likelihood; the posterior is weakly informed in that direction. In this eight-site fit, the strongest remaining boundary issues are the detection and clinical immunity half-saturation terms, the Hill exponents for duration and protection, and the weakly identified infectiousness MOI coefficient $a_m^{\mathrm{inf}}$. The completed NUTS run therefore supports the posterior-predictive fit while preserving honest uncertainty in weak mechanistic directions.

Table 1: **Core response-function equations used by the compartmental model.** State variables are biting-risk stratum $r$, age $a$, immunity $u$, and multiplicity of infection $k$. The fitted parameters column lists the calibrated quantities entering each response function; later tables give code names, priors, and posterior intervals.

| Component | Equation | Fitted parameters | Term definitions |
|---|---|---|---|
| Human force of infection | $\lambda_{r,a,u}(t) = \lambda_0(t)\, m_r\, \frac{\alpha_a}{\bar{\alpha}}\, [1 - p(u)]\, q_a(t)$ | site EIR scales $\log \kappa_s$ | $r$: biting-risk stratum; $a$: age bin; $u$: immunity bin. $m_r$: biting multiplier; $\alpha_a/\bar{\alpha}$: normalized age-biting weight; $q_a(t)$: SMC/vaccine susceptibility multiplier. |
| Infection duration | $D(u) = \mathrm{DUR}_{\text{naive}} - \left(\mathrm{DUR}_{\text{naive}} - \mathrm{DUR}_{\text{floor}}\right) \frac{u^{n_{\text{dur}}}}{u^{n_{\text{dur}}} + (u_{50}^{\text{dur}})^{n_{\text{dur}}}}$ | $\mathrm{DUR}_{\text{naive}}$, $\mathrm{DUR}_{\text{ratio}}$, $u_{50}^{\text{dur}}$, $n_{\text{dur}}$ | $\mathrm{DUR}_{\text{floor}} = \mathrm{DUR}_{\text{naive}}\, \mathrm{DUR}_{\text{ratio}}$; $u_{50}$ gives half-saturation; $n$ gives Hill steepness. |
| Pre-erythrocytic protection | $p(u) = \mathrm{PROT}_{\max} \frac{u^{n_{\text{prot}}}}{u^{n_{\text{prot}}} + (u_{50}^{\text{prot}})^{n_{\text{prot}}}}$ | $\mathrm{PROT}_{\max}$, $u_{50}^{\text{prot}}$, $n_{\text{prot}}$ | $\mathrm{PROT}_{\max}$ is the asymptote; $u_{50}$ and $n$ set midpoint and steepness. |
| Immunity dynamics | $\omega_b(u,k) = b_0(1-u)\log(1+k), \qquad \omega_{\text{wane}}(u, k=0) = \omega_u u$ | $b_0$, $\omega_u$ | $\omega_b$: immunity build; $\omega_{\text{wane}}$: waning in uninfected cells. |
| RDT positivity | $\operatorname{logit} p_{\text{det}}(u,k,a) =$ $d_0 + a_m^{\text{det}} \log(1+k) - \beta_{du} \frac{u}{u + u_{50}^{\text{det}}(a)} - \beta_a g_d(a) - d_{\text{mat}} e^{-a/\tau_{\text{mat}}}$ | $d_0$, $a_m^{\text{det}}$, $\beta_{du}$, $u_{50}^{\text{det}}$, $u_{50}^{\text{det,young}}$, $\beta_a$, $d_{\text{mat}}$ | $g_d(a) = 1 - e^{-a/5}$ is the detection age kernel; $u_{50}^{\text{det}}(a)$ interpolates young and adult half-saturation; $\tau_{\text{mat}}$ is fixed. |
| Clinical incidence | $\ell_{\text{clin}}(u,k,a) = c_0 + c_m \log(1+k) - c_u \frac{u}{u + u_{50}^{\text{clin}}(a)} - c_a m(a) - c_{\text{mat}} e^{-a/\tau_{\text{mat}}}$ $h_{\text{clin}}(u,k,a) = \frac{\operatorname{logit}^{-1}\{\ell_{\text{clin}}(u,k,a)\}}{1 + \tau_{\text{ref}} \operatorname{logit}^{-1}\{\ell_{\text{clin}}(u,k,a)\}}$ | $c_0$, $c_m$, $c_u$, $c_a$, $u_{50}^{\text{clin}}$, $u_{50}^{\text{young}}$, $c_{\text{mat}}$ | $m(a) = 1 - e^{-(a-1)_+/5}$ is the delayed clinical age kernel; $\tau_{\text{ref}} = 14$ d converts raw probability to an event hazard. |
| Human infectiousness | $\operatorname{logit} c(u,k,a) =$ $i_0 + a_m^{\text{inf}} \log(1+k) - \beta_u^{\text{inf}} \frac{u}{u + u_{50}^{\text{inf}}(a)} - \beta_a^{\text{inf}} m(a) - i_{\text{mat}} e^{-a/\tau_{\text{mat}}}$ | $i_0$, $a_m^{\text{inf}}$, $\beta_u^{\text{inf}}$, $\beta_a^{\text{inf}}$, $u_{50}^{\text{inf}}$, $u_{50}^{\text{inf,young}}$, $i_{\text{mat}}$ | $c(u,k,a)$: human infectiousness to mosquitoes; $m(a)$ is the delayed age kernel above; $u_{50}^{\text{inf}}(a)$ interpolates young and adult half-saturation. |
| Population observables | $\mu_{s,a} = \sum_r w_r \frac{\sum_{u,k} P_{s,r,a,u,k}\, p_{\text{det}}(u,k,a)}{\sum_{u,k} P_{s,r,a,u,k}}$ $\lambda_{s,a}^c = 365 \sum_r w_r \frac{\sum_{u,k} P_{s,r,a,u,k}\, h_{\text{clin}}(u,k,a)}{\sum_{u,k} P_{s,r,a,u,k}}$ | none additional | $P_{s,r,a,u,k}$: population state at site $s$; $w_r$: risk-stratum weight; $\mu$: prevalence; $\lambda^c$: annual clinical incidence, not the force of infection. |

Table 2: **Calibrated parameter index for the eight-site NUTS posterior.** Bounds and priors are the bounded natural-scale quantities used in the NUTS target; priors are truncated Gaussians reported as $\mathcal{N}(\mu, \sigma)$ before truncation. Posterior intervals are marginal 95 % credible intervals from the completed NUTS posterior. The per-site median EIR annotations are model-inferred posterior values (base EIR $\times$ exp of the calibrated log-scale) and may differ from the literature nominals cited in the main text.

| Component | Symbol | Code name | Bounds | Prior | Post. 95 % CI | Meaning |
|---|---|---|---|---|---|---|
| Recovery | $\mathrm{DUR}_{\text{naive}}$ | `dur_naive` | $[100, 600]$ d | $(210, 180)$ | $[222, 578]$ d | duration at $u = 0$ |
| Recovery | $\mathrm{DUR}_{\text{ratio}}$ | `dur_ratio` | $[0.05, 0.95]$ | $(0.30, 0.20)$ | $[0.111, 0.779]$ | $\mathrm{DUR}_{\text{floor}}/\mathrm{DUR}_{\text{naive}}$ |
| Recovery | $u_{50}^{\text{dur}}$ | `u50_dur` | $[0.05, 1.5]$ | $(0.40, 0.35)$ | $[0.245, 1.20]$ | immunity half-saturation |
| Recovery | $n_{\text{dur}}$ | `n_dur` | $[1, 5]$ | $(2.0, 3.0)$ | $[1.20, 4.93]$ | Hill exponent |
| Protection | $\mathrm{PROT}_{\max}$ | `prot_max` | $[0.05, 0.95]$ | $(0.30, 0.30)$ | $[0.063, 0.805]$ | maximum pre-erythrocytic protection |
| Protection | $u_{50}^{\text{prot}}$ | `u50_prot` | $[0.05, 1.5]$ | $(0.40, 0.35)$ | $[0.344, 1.28]$ | immunity half-saturation |
| Protection | $n_{\text{prot}}$ | `n_prot` | $[1, 5]$ | $(2.0, 4.2)$ | $[1.52, 4.94]$ | Hill exponent |
| Immunity dynamics | $b_0$ | `b0_scale` | $[0.02, 5.0]$ | $(0.75, 0.90)$ | $[0.097, 0.231]$ | build-rate multiplier |
| Immunity dynamics | $\omega_u$ | `omega_u` | $[0, 1.37\cdot 10^{-3}]$/d | $(2.74\cdot 10^{-4}, 4.93\cdot 10^{-4})$ | $[1.5\cdot 10^{-5}, 1.11\cdot 10^{-3}]$/d | linear waning rate |
| RDT positivity | $d_0$ | `d0_det` | $[-6, 4]$ | $(0.3, 2.5)$ | $[-2.87, -1.01]$ | logit intercept |
| RDT positivity | $a_m^{\text{det}}$ | `a_m_det` | $[0.1, 5]$ | $(1.0, 1.25)$ | $[1.69, 2.68]$ | MOI coefficient on $\log(1 + k)$ |
| RDT positivity | $\beta_{du}$ | `beta_det_u` | $[0.5, 8]$ | $(2.0, 2.55)$ | $[3.19, 5.12]$ | immunity suppression |
| RDT positivity | $u_{50}^{\text{det}}$ | `u50_det` | $[0.05, 1.5]$ | $(0.40, 0.30)$ | $[0.051, 0.193]$ | adult half-saturation |
| RDT positivity | $u_{50}^{\text{det,young}}$ | `u50_det_young` | $[0.10, 1.5]$ | $(0.40, 0.35)$ | $[0.781, 1.48]$ | infant half-saturation |
| RDT positivity | $\beta_a$ | `beta_a` | $[0, 3]$ | $(0.80, 0.45)$ | $[0.641, 0.862]$ | age suppression, shared with clinical |
| RDT positivity | $d_{\text{mat}}$ | `d_mat` | $[0, 5]$ | $(2.0, 3.0)$ | $[0.175, 4.81]$ | maternal-protection magnitude |
| Clinical incidence | $c_0$ | `c0_clin` | $[-6, 2]$ | $(-3.0, 1.0)$ | $[-5.09, -3.80]$ | logit intercept |
| Clinical incidence | $c_m$ | `c_m_clin` | $[0.1, 5]$ | $(1.0, 0.5)$ | $[0.134, 0.790]$ | MOI coefficient |
| Clinical incidence | $c_u$ | `c_u_clin` | $[0.5, 8]$ | $(2.0, 1.5)$ | $[2.25, 5.66]$ | immunity suppression |
| Clinical incidence | $c_a$ | `c_a_clin` | $[0, 4]$ | $(2.0, 1.0)$ | $[1.28, 2.79]$ | age suppression |
| Clinical incidence | $u_{50}^{\text{clin}}$ | `u50_clin` | $[0.05, 1.5]$ | $(0.40, 0.20)$ | $[0.057, 0.580]$ | adult half-saturation |
| Clinical incidence | $u_{50}^{\text{young}}$ | `u50_young` | $[0.05, 1.5]$ | $(0.40, 0.20)$ | $[0.110, 0.828]$ | infant half-saturation, shared with detection |
| Clinical incidence | $c_{\text{mat}}$ | `c_mat` | $[0, 5]$ | $(2.0, 2.0)$ | $[0.293, 4.81]$ | maternal-protection magnitude |
| Infectiousness | $i_0$ | `i0_inf` | $[-6, 0]$ | $(-3.0, 0.8)$ | $[-4.51, -3.01]$ | logit intercept |
| Infectiousness | $a_m^{\text{inf}}$ | `a_m_inf` | $[0, 4]$ | $(1.0, 0.6)$ | $[0.007, 0.699]$ | MOI coefficient on $\log(1 + k)$ |
| Infectiousness | $\beta_u^{\text{inf}}$ | `beta_inf_u` | $[0, 8]$ | $(3.0, 1.5)$ | $[1.48, 4.74]$ | immunity suppression |
| Infectiousness | $\beta_a^{\text{inf}}$ | `beta_a_inf` | $[0, 4]$ | $(1.0, 0.8)$ | $[0.142, 1.57]$ | age suppression |
| Infectiousness | $u_{50}^{\text{inf}}$ | `u50_inf` | $[0.05, 0.95]$ | $(0.30, 0.15)$ | $[0.104, 0.595]$ | adult half-saturation |
| Infectiousness | $u_{50}^{\text{inf,young}}$ | `u50_inf_young` | $[0.05, 0.95]$ | $(0.60, 0.25)$ | $[0.143, 0.921]$ | infant half-saturation |
| Infectiousness | $i_{\text{mat}}$ | `i_mat` | $[0, 5]$ | $(2.0, 1.5)$ | $[0.196, 4.70]$ | maternal-protection magnitude |
| Site EIR scale | $\log \kappa_s$ | `log_eir_scale_0` | $[-0.7, 0.7]$ | $(0.0, 0.2)$ | $[-0.489, +0.208]$ | Namawala 1991; median EIR $\approx 283$ |
| Site EIR scale | $\log \kappa_s$ | `log_eir_scale_1` | $[-0.7, 0.7]$ | $(0.0, 0.2)$ | $[-0.651, -0.146]$ | Ndiop 1993; median EIR $\approx 13$ |
| Site EIR scale | $\log \kappa_s$ | `log_eir_scale_2` | $[-0.7, 0.7]$ | $(0.0, 0.2)$ | $[-0.230, +0.355]$ | Dielmo 1990; median EIR $\approx 211$ |
| Site EIR scale | $\log \kappa_s$ | `log_eir_scale_3` | $[-0.7, 0.7]$ | $(0.0, 0.2)$ | $[-0.586, +0.012]$ | Chonyi 1999; median EIR $\approx 26$ |
| Site EIR scale | $\log \kappa_s$ | `log_eir_scale_4` | $[-0.7, 0.7]$ | $(0.0, 0.2)$ | $[+0.159, +0.675]$ | Ngerenya 1999; median EIR $\approx 6$ |
| Site EIR scale | $\log \kappa_s$ | `log_eir_garki_0` | $[-1.0, 1.0]$ | $(0.0, 0.28)$ | $[-0.293, +0.393]$ | Matsari; median EIR $\approx 66$ |
| Site EIR scale | $\log \kappa_s$ | `log_eir_garki_1` | $[-1.0, 1.0]$ | $(0.0, 0.28)$ | $[+0.350, +0.906]$ | Rafin Marke; median EIR $\approx 34$ |
| Site EIR scale | $\log \kappa_s$ | `log_eir_garki_2` | $[-1.0, 1.0]$ | $(0.0, 0.28)$ | $[-0.944, -0.319]$ | Sugungum; median EIR $\approx 69$ |

Recovery / infection duration follows the Hill S-curve in Table 1.

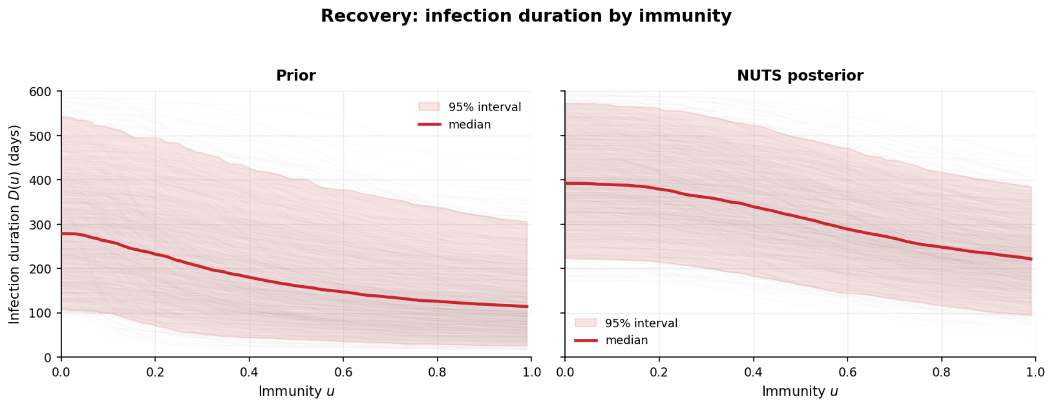


Figure 1: **Recovery:** $D(u)$ **under the prior (left) vs. posterior (right) under the current eight-site NUTS posterior.** 1000 prior draws (independent truncated Gaussians from Table 2) and 1000 posterior draws subsampled from the 2160-draw NUTS posterior are each pushed through the Hill S-curve. Red line is the median; pink band is the pointwise 95 % interval. The NUTS posterior narrows the duration curve relative to the prior but still leaves substantial uncertainty in the naive duration, floor ratio, and Hill exponent.

Pre-erythrocytic protection follows the Hill S-curve in Table 1.

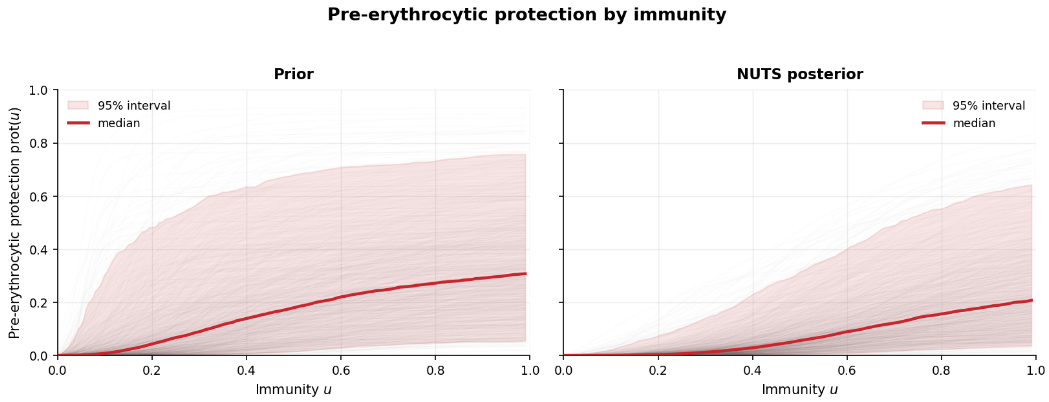


Figure 2: **Protection:** $\text{PROT}(u)$ **under the prior (left) vs. posterior (right) under the current eight-site NUTS posterior.** 1000 prior draws (independent truncated Gaussians from Table 2) and 1000 posterior draws subsampled from the 2160-draw NUTS posterior are pushed through the protection Hill curve. $\text{PROT}_{\text{max}}$ is centered near $0.33$ but remains weakly constrained, and $n_{\text{prot}}$ retains broad support up to the upper bound.

Immunity build is driven by exposure with MOI log-saturation, and waning is linear on uninfected cells only (Table 1).

Because $u$ is a latent exposure-history coordinate rather than a directly measured immunological quantity, the absolute build and waning rates should be interpreted jointly with the fitted response curves for infection duration, protection, detection, clinical incidence, and infectiousness. Slow movement along the $u$ axis can still produce substantial epidemiological effects when the fitted response curves are steep at low $u$.

RDT detection is modelled as a logistic regression in immunity, MOI, age, and a maternal-protection floor (Table 1), where $g_d(a) = 1 - \exp(-a/5)$. The half-saturation point $u_{50}^{\text{det}}(a)$ smoothly interpo-

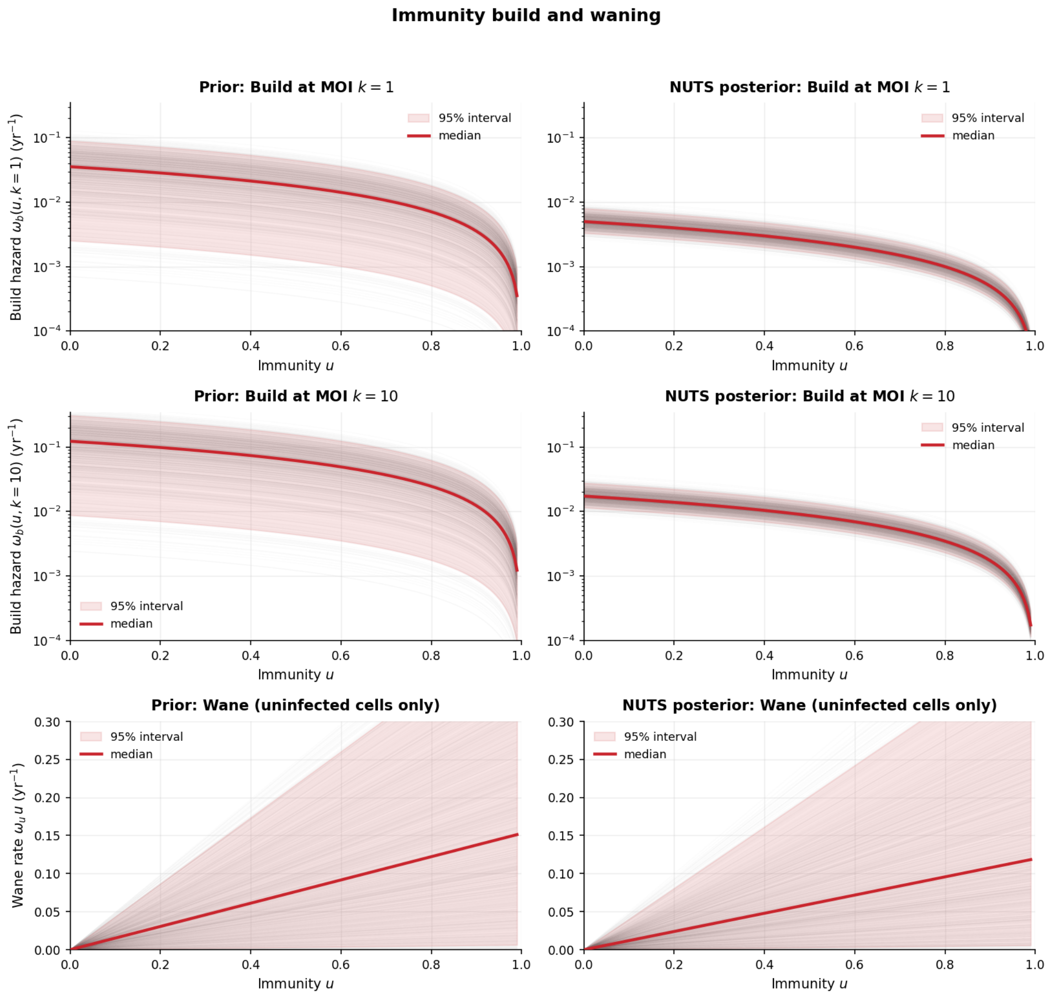


Figure 3: **Immunity dynamics: build hazard at low and high MOI (top and middle) and wane rate (bottom), under the prior (left) vs. posterior (right) under the current eight-site NUTS posterior.** Rates are converted to per-year units for readability ($\times 365$). Build is evaluated at MOI $k = 1$ and $k = 10$ to show the $\log(1 + k)$ scaling. The build panels use logarithmic $y$-axes; the wane panels remain linear. Posterior build rates are slower than the prior expects (median $b_0 \approx 0.15$ vs. prior centre $0.75$). Waning remains weakly constrained but is concentrated near small values, consistent with slow loss of immunity.

lates from $u_{50}^{\text{det,young}}$ in infants to $u_{50}^{\text{det}}$ in adults using $s(a) = 1 - \exp[-(a-1)_+/5]$. Detection is set to zero for uninfected cells ($k = 0$). Site/age RDT prevalence is the risk-weighted compartmental average in Table 1.

Clinical incidence uses the logistic structure in Table 1, but with an age kernel that switches on at age 1 yr rather than from birth: $m(a) = 1 - e^{-(a-1)_+/5}$. This is the same kernel that interpolates $u_{50}^{\text{clin}}(a)$ between $u_{50}^{\text{young}}$ (infants) and $u_{50}^{\text{clin}}$ (adults). The maternal-protection kernel is shared in form with detection; $u_{50}^{\text{young}}$ is shared with detection's young-children half-saturation. The raw clinical probability is $p_{\text{clin}} = \text{logit}^{-1}(\ell_{\text{clin}})$ for infected cells and zero when $k = 0$. To avoid counting repeated clinical days within the same infectious episode as independent episodes, the raw probability is converted to the effective per-day hazard $h_{\text{clin}}$ in Table 1. Annual site/age incidence is then the risk-weighted compartmental average in the same table.

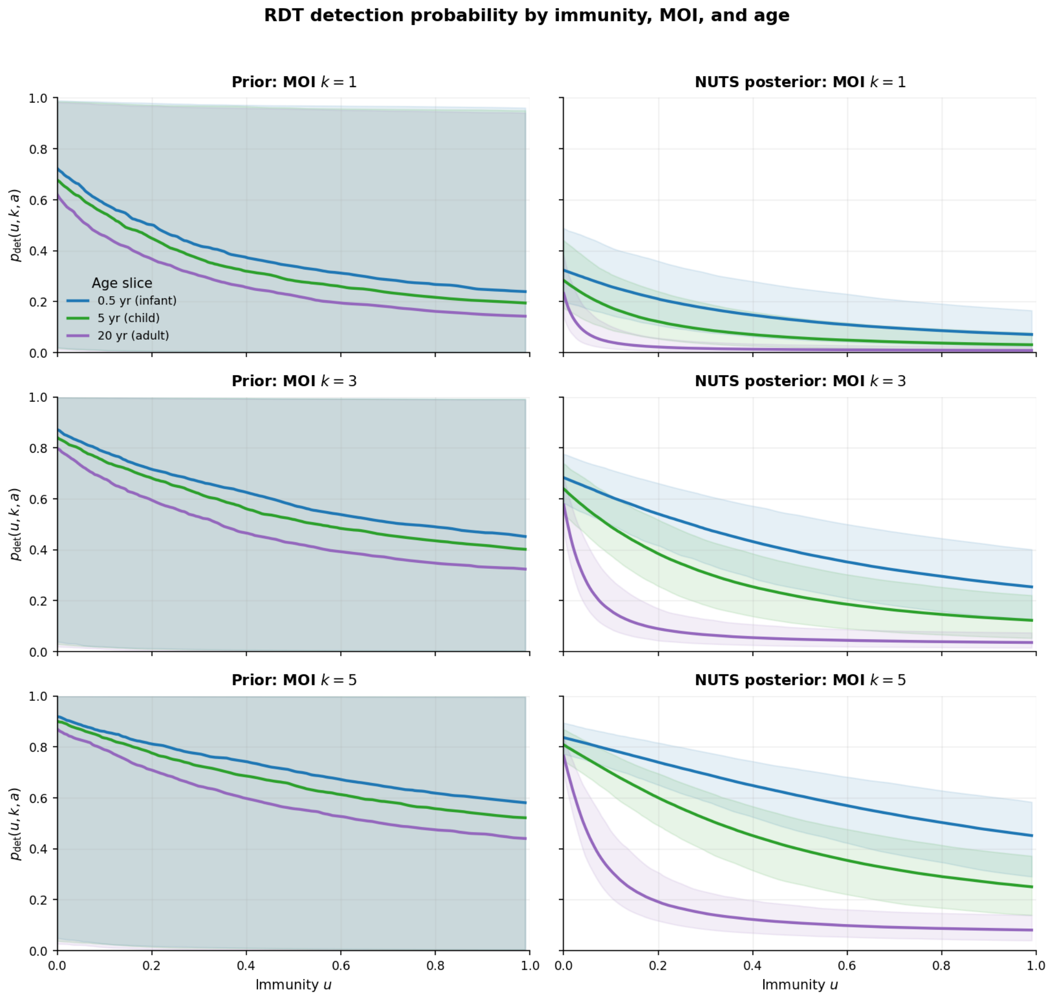


Figure 4: **Detection: $p_{\mathbf{det}}(u,k,a)$ under the prior (left) vs. posterior (right).** Rows are MOI levels ($k = 1, 3, 5$); colours are age slices (0.5, 5, and 20 yr). Lines and bands show medians and pointwise 95 % intervals from 1000 draws. The posterior tightens the MOI increase and immunity-driven decline in RDT positivity.

Infectiousness $c(u, k, a)$ is calibrated in the eight-site posterior. The functional form mirrors detection and clinical: the logistic equation in Table 1, where $u_{50}^{\mathrm{inf}}(a)$ interpolates from $u_{50}^{\mathrm{inf,young}}$ (infants) to $u_{50}^{\mathrm{inf}}$ (adults) via the same $m(a)$ kernel used for clinical incidence. The 7 kernel parameters are calibrated to gametocyte-density-derived human-to-mosquito infectivity per (site, age bin, survey) at the three Garki villages via the Churcher et al. 2013 saturating curve [Churcher et al., 2013] (Appendix M, data-aggregation subsection).

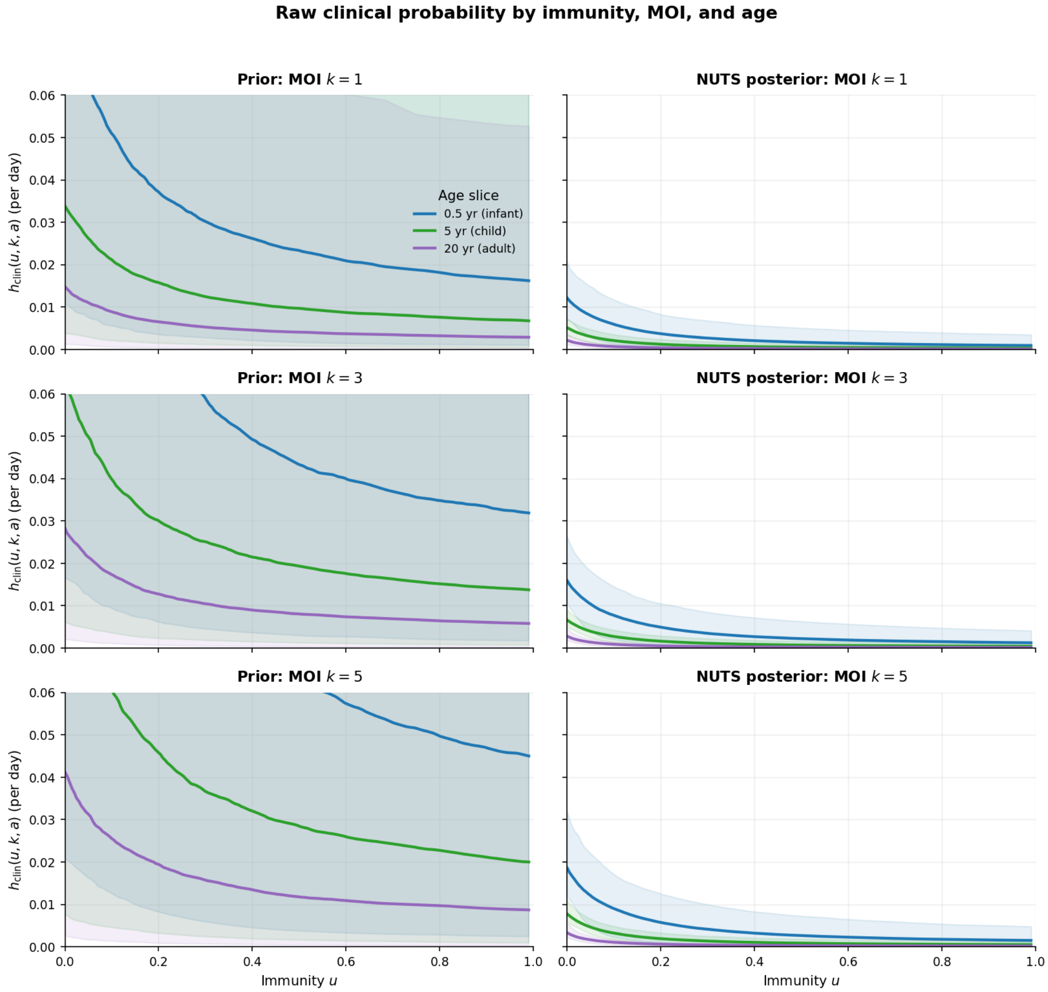


Figure 5: **Raw clinical probability $p_{\text{clin}}(u, k, a)$ under the prior (left) vs. posterior (right).** Rows are MOI levels ($k = 1, 3, 5$); colours are age slices (0.5, 5, and 20 yr). Lines and bands show medians and pointwise 95 % intervals from 1000 draws, evaluated before the refractory adjustment. The posterior strongly narrows the clinical-probability surface.

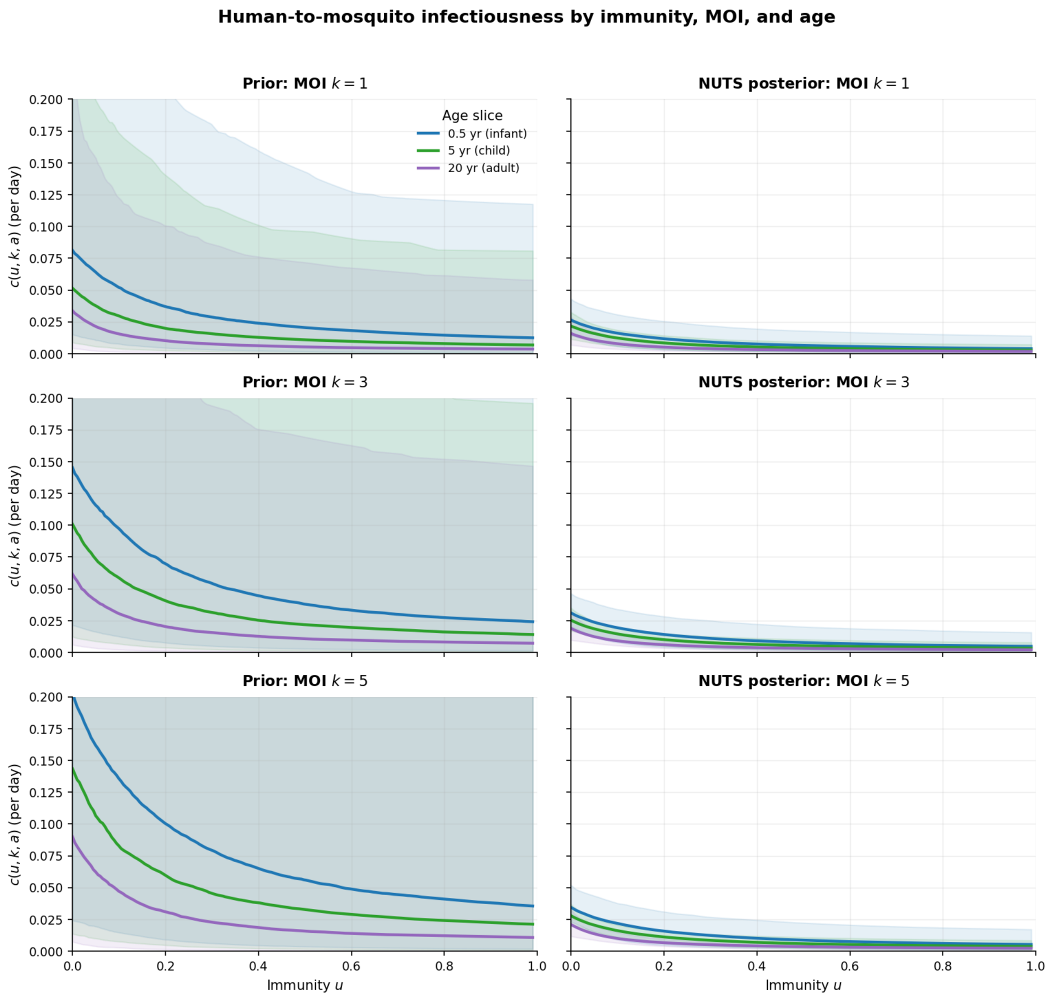


Figure 6: **Infectiousness $c(u, k, a)$ under the prior (left column) vs. posterior (right column) under the current eight-site NUTS posterior, at three MOI levels (rows: $k = 1, 3, 5$) and three age slices per panel.** 1000 prior draws and 1000 posterior draws subsampled from the 2160-draw NUTS posterior are pushed through the logistic kernel. The Garki infectiousness targets identify the broad scale and immunity/age suppression, while several shape terms remain weakly constrained, especially the MOI coefficient $a_m^{\text{inf}}$.

#### Fixed structural constants and response kernels

Most natural-history response functions are estimated in the eight-site posterior: recovery, protection, RDT positivity, clinical incidence, human infectiousness, and site EIR scales are the calibrated components listed in Tables 1 and 2. A smaller set of quantities remains fixed because it defines the discretisation, ageing structure, age-biting and age-effect kernels, observation timing, and base scaling constants that are not identifiable from the population-level calibration data (Table 3).

Table 3: **Fixed structural constants, response-function kernels, and vector priors.** None are estimated in the eight-site posterior; the calibrated parameters are in Tables 1 and 2. Vector-biology priors marked *intv only* are sampled in the intervention/decision sweep but held at their medians during calibration.

| Quantity | Value | Notes |
|---|---|---|
| Age bin edges (yr) | $[0, 1, 2, 5, 10, 15, 20, 30, 40, 50, 60, 75]$ | 11 bins (12 edges); infant resolution |
| Immunity bins $n_u$ | 10 | uniform on $[0, 0.99]$ |
| MOI cap $K_{\max}$ | 15 | 16 MOI levels total |
| Risk strata | 5 strata, equal weights | mults $[0.107, 0.36, 0.70, 1.22, 2.61]$ |
| Birth rate | 35.25/1000/yr | high-fertility stable-population driver |
| Age-death shape | 12 anchor points | mortality multipliers rescaled so births = deaths |
| Stable age distribution | 14.8% $<$ 5 yr; 41.8% $<$ 15 yr | implied by birth rate, age bins, and rescaled death shape |
| Age-biting weights $\alpha_a$ | surface-area age-biting curve | continuous, bin-averaged; approximates surface-area dependence [Gerardin et al., 2015] |
| Time step $\Delta t$ | 1 d | exponential Euler with mass correction |
| $\tau_{\text{mat}}$ | 0.2055 yr ($\sim$ 75 d) | shared by detection & clinical |
| Age maturation kernels | $g_d(a)$; $m(a)$ | detection vs. clinical/infectiousness age effects |
| $\tau_{\text{ref}}$ | 14 d | clinical refractory period |
| Immunity build scale | $b_{0,\text{base}} = 1.35{\times}10^{-4}$/d, $\gamma_b = 1.0$, $k_b = 3.0$ | base rate, immunity exponent, MOI saturation; the $b_0$ multiplier is calibrated |
| EIP prior | LogN med. 11 d, $[8, 15]$ | *intv only* |
| $V2H$ prior | TN med. 0.50, $[0.20, 0.80]$ | vector-to-human prob, *intv only* |
| ITN usage prior | TN med. 0.85, $[0.65, 0.95]$ | retention-conditional, *intv only* |

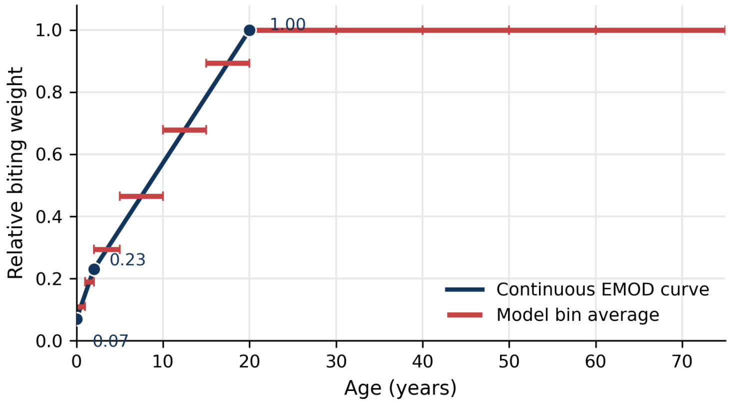


Figure 7: **Surface-area age-biting weight.** The model uses a piecewise surface-area age-biting curve (blue), bin-averaged onto the compartmental age grid (red). Gerardin et al. [2015] describe using age-dependent biting risk to approximate surface-area dependence.

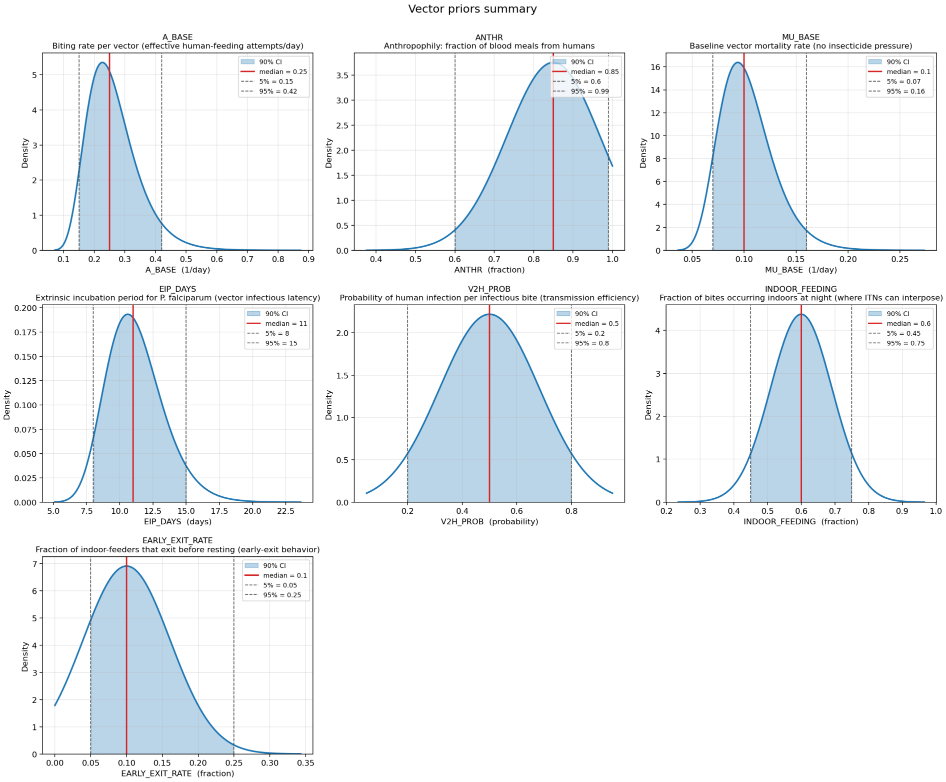


Figure 8: **Vector-biology priors used in matched-EIR decision projections.** Marginal prior densities for the seven vector parameters sampled in the vector-uncertainty layer. Blue shading denotes the central 90% prior interval and red vertical lines mark medians. These priors are not updated by the eight-site calibration; matched-EIR conditioning adjusts $K_{\text{wet}}$ so each retained draw is compared at the target annual EIR.

### B Differentiable implementation details

This appendix explains how DELENDA differentiates decades-long simulations at daily timesteps without storing an impractically large computation graph.

The dominant computational cost in DELENDA is the human–vector ODE solve. A calibration forward pass integrates 60 years of daily steps over an 8,804-dimensional state for each of the 8 calibration sites. Naively differentiating this calculation with `jax.grad` would retain the full daily trajectory and all autodiff intermediates, exceeding single-GPU memory for the posterior and decision-analysis workflows.

We therefore wrap the ODE solver in a custom vector-Jacobian product. The forward pass runs the same exponential-Euler-with-mass-correction solver used elsewhere in the manuscript, but stores only segment-end states. During the backward pass, the solver recomputes the daily steps within each segment while propagating adjoint sensitivities in reverse. The memory cost is therefore $O(\text{state size} \times n_{\text{segments}})$ rather than $O(\text{state size} \times n_{\text{steps}})$, with wall-clock cost approximately twice a forward pass. Once wrapped this way, the solver is a normal JAX primitive: it can be JIT compiled, vmapped across sites and posterior draws, and differentiated through by posterior calibration and intervention-mix optimization.

### C Matched-EIR implementation details

This appendix defines the conditioning step used to compare uncertain posterior/vector draws at the same annual EIR.

For each posterior/vector draw, DELENDA solves for the wet-season larval-recruitment amplitude $K_{\text{wet}}$ that produces the target annual EIR while holding the seasonal phase and dry-season floor fixed:

$$K_L(t) = K_{\text{dry}} + K_{\text{wet}} s_0(t), \qquad s_0(t) = \frac{s(t) - \min_t s(t)}{\max_t s(t) - \min_t s(t)}, \quad K_{\text{dry}} = 0.10.$$

Here $s(t)$ is the normalized monthly seasonality profile. This conditioning step makes the EIR label refer to the same baseline transmission intensity across posterior and vector-biology draws, rather than allowing uncertainty in mosquito traits to also move the achieved EIR.

Draws that cannot reproduce the requested EIR within the search range are excluded from the matched-EIR conditional analysis. These exclusions are useful prior-predictive diagnostics: they identify vector-prior combinations whose vectorial capacity is incompatible with the target EIR. Retention exceeds 99% across the decision grid, with the only material exclusions at the lowest target EIR.

#### D Illustrative Dapelogo-inspired routine-care baseline

This appendix provides a scale check for the decision-analysis outcomes by showing the routine-care burden implied by the matched-EIR Dapelogo-inspired setup. The decision-analysis figures optimize added prevention interventions against a matched-EIR Dapelogo-inspired baseline with fixed routine case management; to make that baseline concrete, Figures 9 and 10 show the routine-care clinical burden and seasonality used to orient the decision sweeps. The figures condition on annual EIR rather than latent larval carrying capacity: $K_{\text{wet}}$ is matched to the underlying transmission potential before routine case management, then held fixed while routine treatment is enabled.

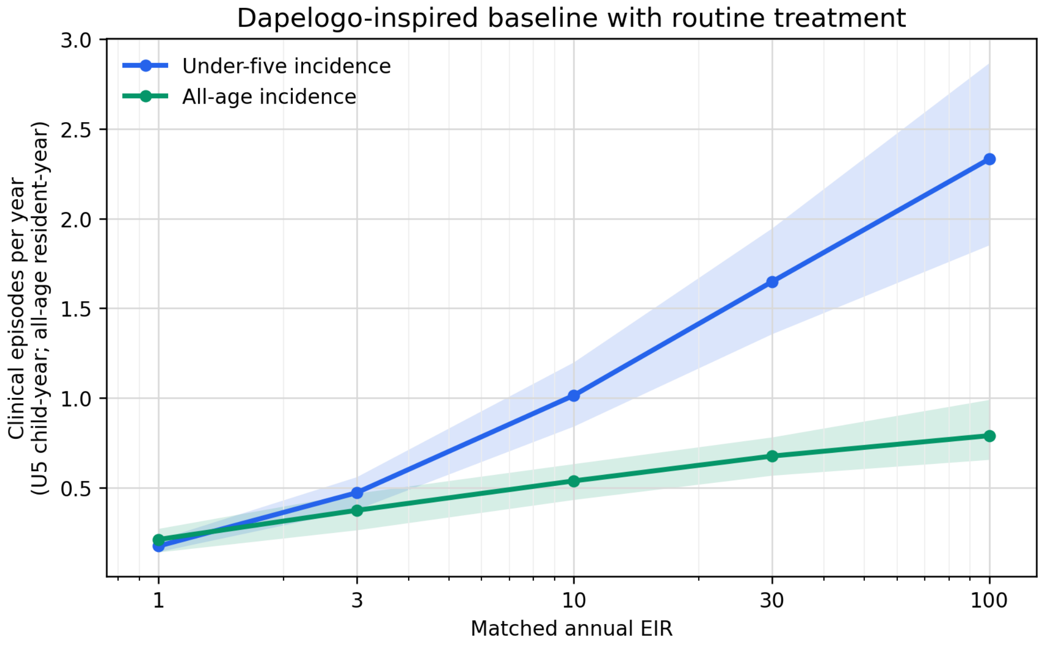


Figure 9: **Routine-care clinical burden across matched EIR.** Posterior and vector-biology uncertainty are propagated through the Dapelogo-inspired matched-EIR baseline used in the decision analysis, with the four optimized prevention interventions set to zero and fixed routine treatment maintained. Lines show median annualized clinical incidence for under-five children and all ages; shaded bands show central 95% intervals across retained matched-EIR draws. Empirical calibration data are not overlaid here because the purpose is to orient the illustrative decision baseline rather than to repeat the posterior-predictive fit in the main text.

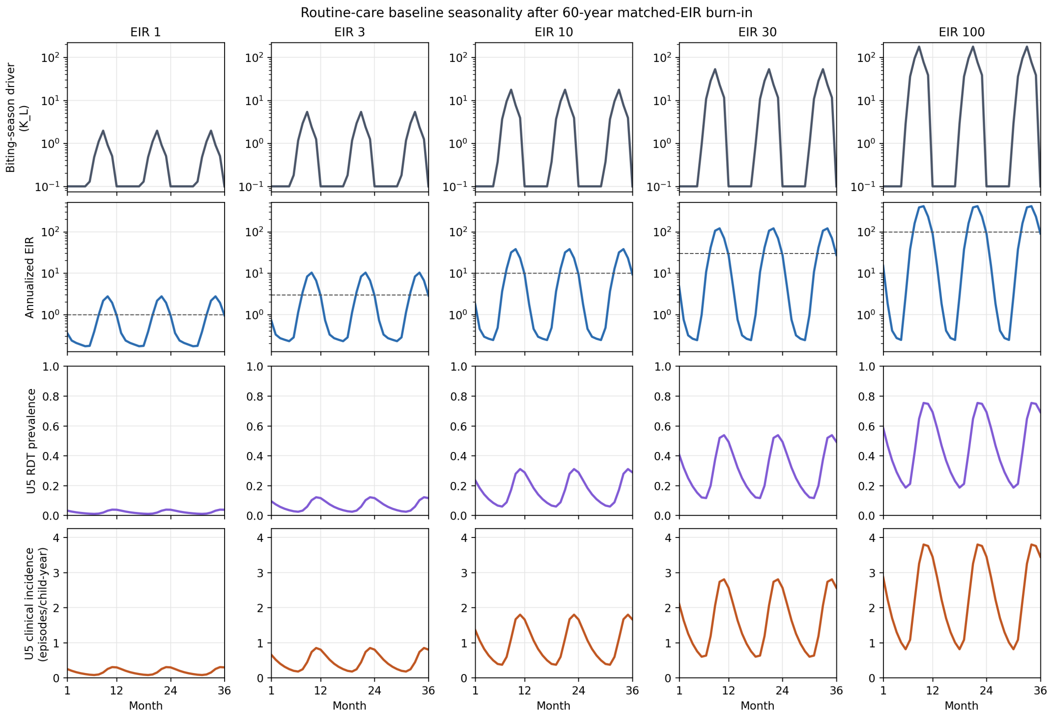


Figure 10: **Routine-care seasonality across matched EIR.** Baseline dynamics after a 60-year burn-in for the five EIR levels used in the decision grid, with routine treatment maintained and the four optimized prevention interventions set to zero. The fixed dry-season larval-capacity floor keeps the dry season dry across transmission settings, while the wet-season component scales to match the target annual EIR.

### E Intervention cost-prior details

This appendix documents the unit conversions and uncertainty ranges that place intervention-specific programme costs on the common resident-equivalent budget scale.

The optimization cost model uses resident-equivalent costs over the 3-year decision horizon: dollars per resident after any target-population scaling needed to place population-wide interventions (ITNs and IRS) and child-targeted interventions (SMC and vaccination) on a common budget scale. These are not intended as a full accounting model; they are decision-analysis priors. Unit costs use lognormal priors, summarized below by their median and central 90% interval. Convexity coefficients $\gamma_i$ use normal priors with the listed central 90% intervals and are operational-convexity planning priors, not fitted marginal-cost curves. The isolated single-intervention cost-effect curves in Figure 11 provide the visual summary of how these cost priors translate into marginal impact at different coverage levels and EIR settings.

The entries in Table 4 should be read as planning priors rather than local budget estimates. ITN and IRS costs are already population-wide resident-equivalent costs. SMC and vaccination begin from eligible-child programme costs and are placed on the resident-equivalent scale using the model's under-five population fraction, 0.148. The ITN prior assumes one distribution campaign over the 3-year horizon and roughly one delivered net per two residents; IRS assumes annual spraying; SMC assumes one 4-cycle seasonal campaign per year; and the vaccine prior represents a complete protective course for eligible children.

The overhead terms $\gamma_i$ represent increasing marginal delivery effort at high coverage, including household registration, transport, repeat visits, supervision, wastage, caregiver return visits, and reaching harder-to-reach households. They are deliberately simple operational-convexity priors, not fitted marginal-cost curves. The medians and uncertainty ranges are anchored to published malaria

Table 4: Cost-prior medians and uncertainty ranges used in the intervention-mix optimization. Costs are resident-equivalent US dollars over the 3-year decision horizon after target-population scaling. The SMC row shows both the target-child programme cost and the resident-equivalent value used in $C(\boldsymbol{\theta}_{\text{int}})$. The vaccine row is likewise a resident-equivalent programme cost, not a literal per-dose price.

| Tool | Base-cost prior | Full-coverage cost at median overhead | Convexity prior |
|---|---|---|---|
| ITN | LogN median \$1.80 per resident over 3 years; 90% interval \$1.20–\$2.70 | \$2.34 per resident | $\gamma_{\text{ITN}} = 0.30$; 90% interval 0.10–0.50 |
| IRS | LogN median \$22.50 per resident over 3 years; 90% interval \$15–\$35 | \$29.25 per resident | $\gamma_{\text{IRS}} = 0.30$; 90% interval 0.15–0.45 |
| SMC | LogN median \$11.40 per eligible child over 3 annual SMC seasons; 90% interval \$8–\$20; scaled by under-five fraction 0.148 to \$1.69 per resident | \$2.54 per resident; \$17.10 per eligible child | $\gamma_{\text{SMC}} = 0.50$; 90% interval 0.30–0.70 |
| Vaccine | LogN median \$2.70 per resident-equivalent programme course; 90% interval \$1.50–\$5.00 | \$4.59 per resident; about \$31 per fully vaccinated eligible child | $\gamma_{\text{Vacc}} = 0.70$; 90% interval 0.40–1.00 |

intervention delivery-cost syntheses and cost-effectiveness studies [Yukich et al., 2008, Kolaczinski et al., 2010, Smith Paintain et al., 2014, Arroz et al., 2019, Alonso et al., 2021, Yukich et al., 2022, Pitt et al., 2017, Conteh et al., 2021, Winskill et al., 2017, World Health Organization, 2023, Glassman and Vaca, 2021]. Operational applications should replace these rows with local procurement, delivery, wastage, and financing assumptions.

#### F Single-intervention cost-effect diagnostics

Figure 11 isolates each intervention by varying its coverage while holding the other three optimized prevention interventions at zero and maintaining routine treatment. This is not an optimized portfolio analysis: because interactions are absent, it should be read as marginal intuition for the main-text optimized portfolio frontier. The low-cost, steep initial ITN curves explain why ITNs enter first in the optimizer, while IRS shows larger nominal cost and broader uncertainty. SMC and vaccination have larger under-five than all-age effects because their modeled deployment is child-targeted.

Figures 12 and 13 decompose the same isolated single-intervention effects by uncertainty source. The pattern is intervention-specific rather than uniform. Vaccine-effect variance is dominated by efficacy uncertainty, IRS variance is driven largely by vector biology and efficacy, and SMC uncertainty shifts with the objective and transmission setting. These diagnostics are not portfolio optimizations, but they indicate where additional data collection or local elicitation would most improve future decision analyses: product efficacy for vaccine, vector-behavioural inputs for IRS, and posterior/model uncertainty for parts of the SMC response surface.

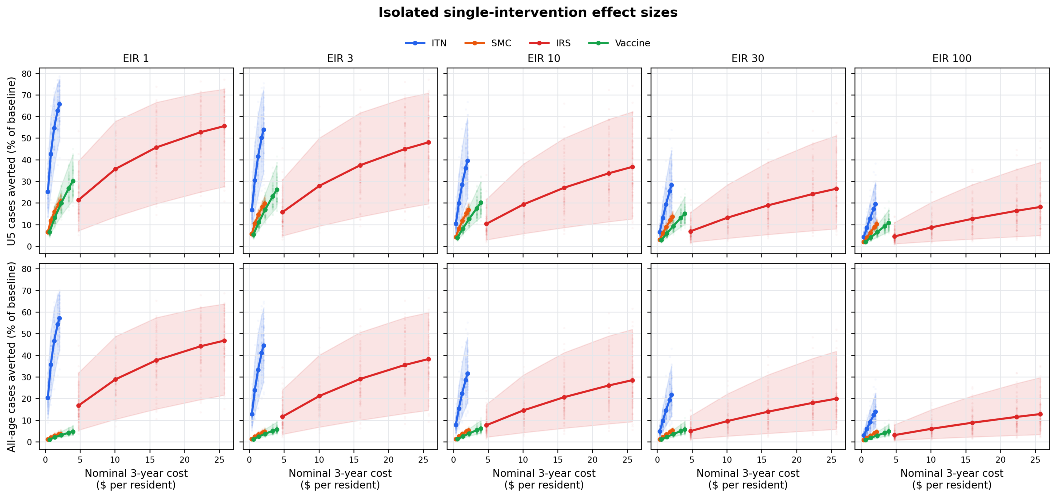


Figure 11: **Isolated single-intervention cost-effect curves.** Each panel fixes matched annual EIR and shows one intervention at a time, with all other optimized prevention interventions held at zero. Routine treatment is maintained. The $x$ axis is the nominal 3-year resident-equivalent cost implied by the median cost coefficients and the selected coverage; it does not include cost-prior uncertainty. Lines show mean cases averted as a percentage of the routine-care baseline, shaded bands show central 95% intervals across posterior, vector, and intervention-efficacy draws, and faint vertical point clouds show individual draw-level outcomes.

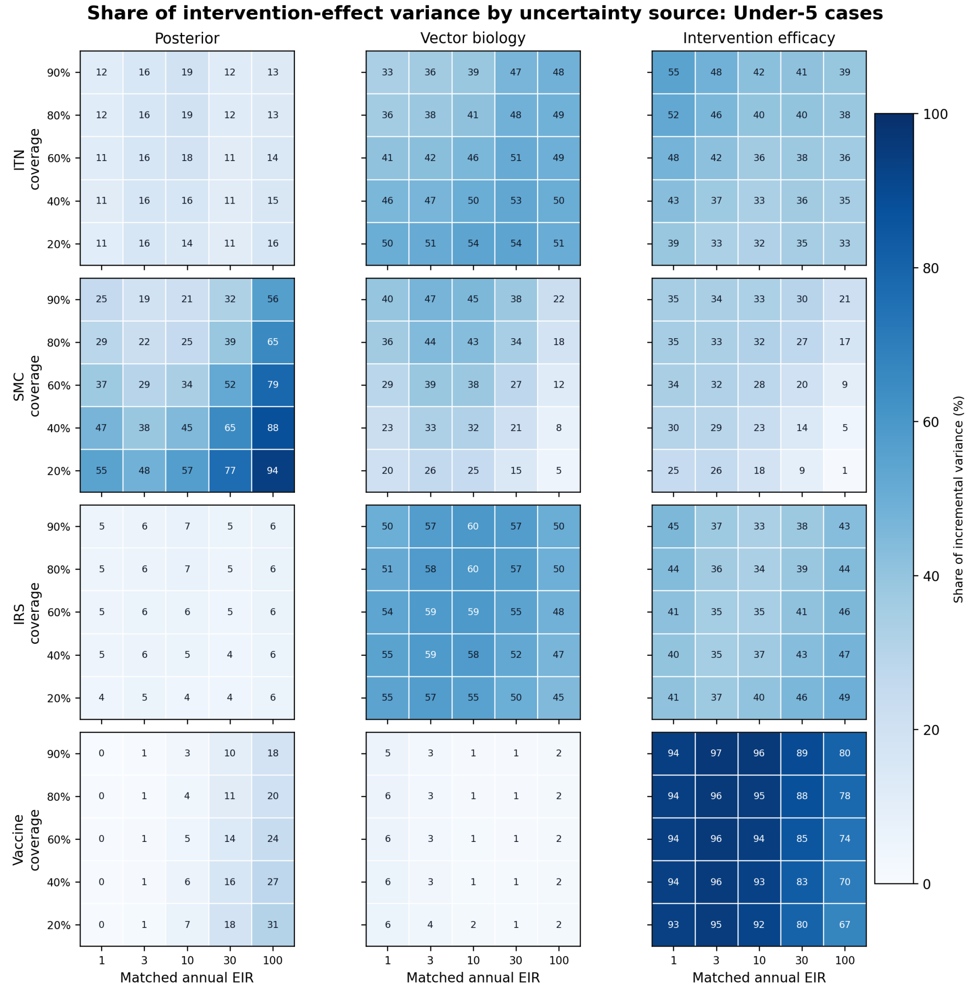


Figure 12: **Uncertainty sources for isolated under-five intervention effects.** Each cell shows the share of incremental variance in predicted under-five cases averted attributable to the calibrated posterior, vector-biology priors, or intervention-efficacy priors in the single-intervention sweep. Rows vary the intervention and coverage level; columns vary the matched annual EIR.

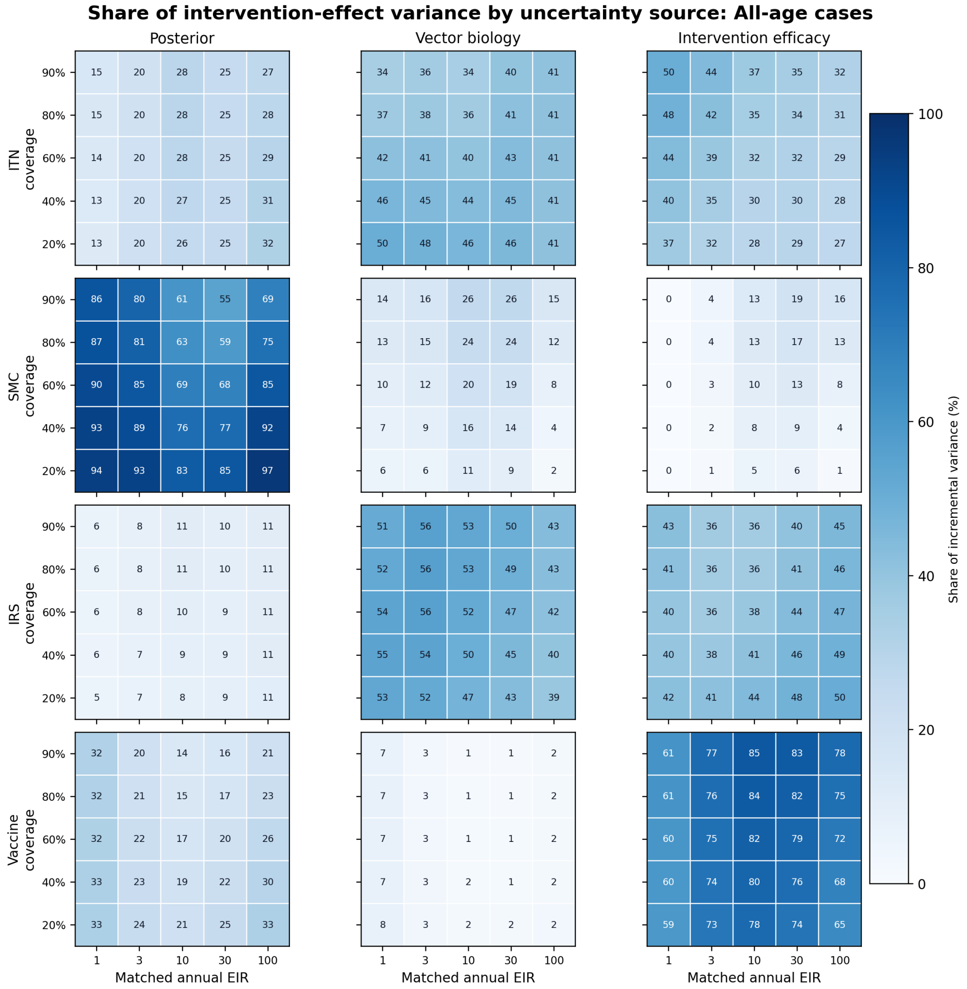


Figure 13: **Uncertainty sources for isolated all-age intervention effects.** Format follows Figure 12, but the outcome is all-age cases averted. Different uncertainty layers matter for different interventions and transmission levels, reinforcing that future uncertainty reduction should be targeted rather than generic.

### G Intervention effect-size consistency checks

Figure 14 is a prior-predictive consistency diagnostic, not a formal fit of intervention-effect parameters to external trials. It asks whether the mechanistic intervention modules, when combined with the calibrated transmission model and independently specified priors, produce effect sizes in the range of empirical evidence. The comparison axis differs by intervention because the most defensible empirical anchor differs: ITN and IRS are shown as coverage-effect curves against pooled coverage evidence; SMC is shown across a model EIR sweep with empirical ranges rather than EIR-positioned trial points; and the R21 vaccine check is shown against observed control-arm clinical incidence to avoid assigning trial sites an unobserved EIR. The panels are retained as qualitative consistency checks for the intervention modules. They are not used as inputs to the decision analysis; the decision figures in the main text come from the NUTS-based optimization runs.

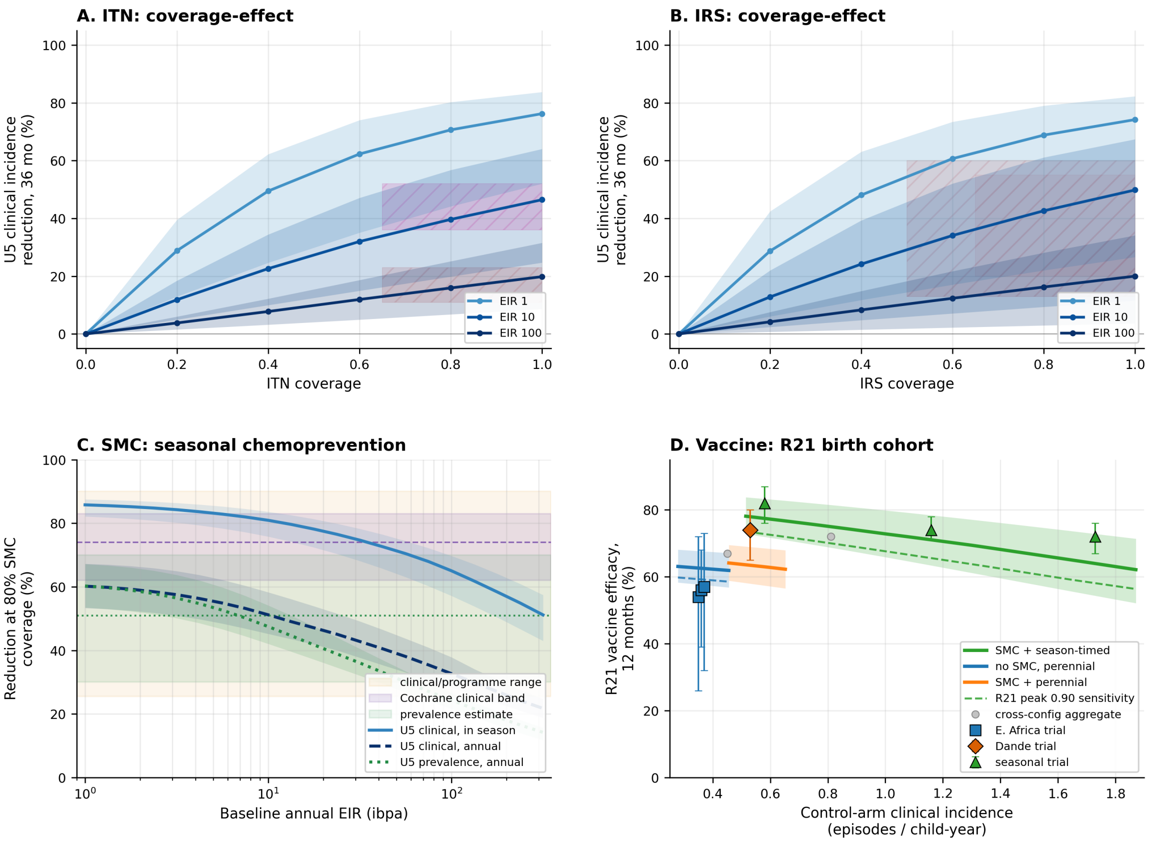


Figure 14: **Cross-intervention prior-predictive effect-size consistency checks.** Panels A and B show 36-month under-five clinical incidence reductions for isolated ITN and IRS deployment across three matched baseline EIRs, with joint model/vector/intervention uncertainty bands and pooled empirical anchors. Panel C shows SMC at 80% coverage across matched annual EIR, comparing in-season clinical-incidence, annual clinical-incidence, and annual parasite-prevalence reductions with empirical evidence ranges. Panel D shows the R21 birth-cohort prior-predictive check against Phase 3 trial sites on the matched control-arm clinical-incidence axis. These panels are qualitative consistency checks; the empirical anchors are not fitted likelihood terms and are not used as inputs to the decision analysis.

### H ITN priors and effect-size check

Panel A of Figure 14 shows isolated ITN coverage–effect curves for under-five clinical incidence. The empirical anchor is the pooled Cochrane evidence for insecticide-treated nets: the clinical-malaria summary from Pryce et al. [2018] (RR $0.55$, 95% CI $0.48$–$0.64$) and the lower, endpoint-disjoint all-cause child-mortality summary (RR $0.83$, 95% CI $0.77$–$0.89$; Lengeler 2004, Pryce et al. 2018). The mortality band is shown only as a lower-effect comparator; it is not converted to clinical incidence.

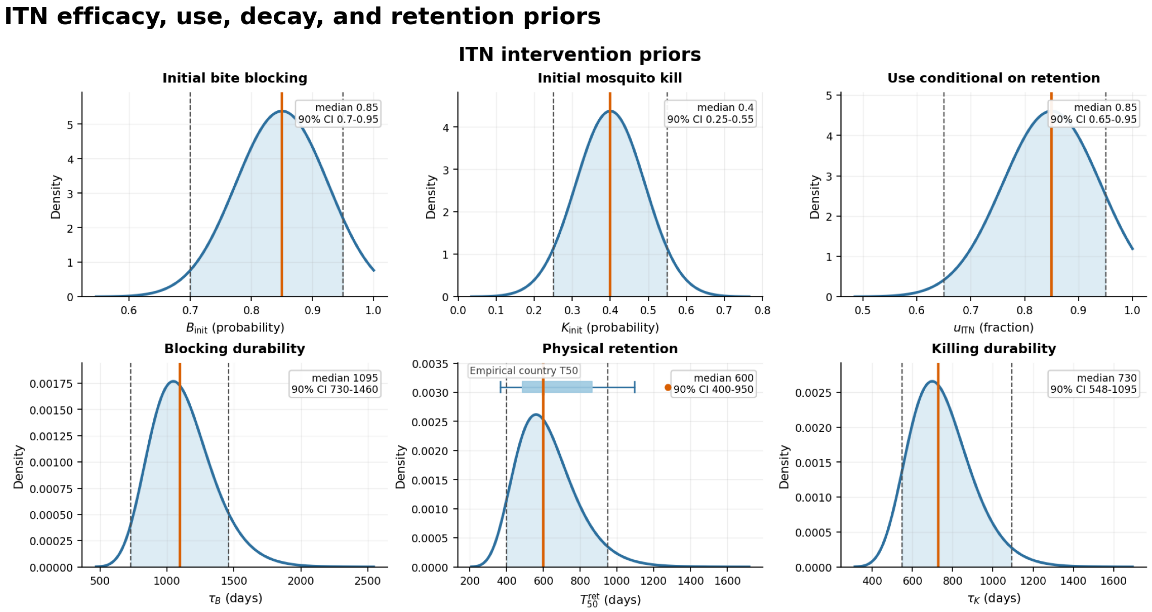


Figure 15: **ITN intervention priors.** The figure shows the six ITN-specific efficacy, usage, decay, and retention priors used in the decision analysis. Indoor-night biting is excluded because it is sampled in the shared vector-behavioural layer used by both ITN and IRS.

The ITN module samples intervention-specific priors for physical blocking, mosquito killing, use conditional on retention, durability of blocking and killing, and physical retention (Figure 15). The shared indoor-night biting fraction is sampled in the vector layer, so ITN and IRS use the same exposure draw. The blocking prior is centered high, the kill prior is deliberately broad and lower to span insecticide resistance and product heterogeneity, and retention follows a Weibull curve anchored to sub-Saharan African net-retention estimates. These are generic ITN planning priors, not a product-specific comparison among pyrethroid-only, PBO, and dual-active-ingredient nets [Killeen et al., 2007, Pryce et al., 2018, Bertozzi-Villa et al., 2021, Mosha et al., 2022, Accrombessi et al., 2023, Skovmand and Lengeler, 2008, Tan et al., 2016, Cooke et al., 2023].

The ITN panel is therefore a qualitative prior-predictive check against pooled evidence, not a new meta-analysis and not a likelihood term. Next-generation-net and net-vs-net trials answer an incremental product-choice question and are outside this generic ITN-vs-control diagnostic.

### I IRS priors and effect-size check

Panel B of Figure 14 shows isolated IRS coverage–effect curves for under-five clinical incidence. The IRS module samples four intervention-specific quantities: fresh-substrate laboratory kill, field attenuation from laboratory to field conditions, operational spray fidelity, and residual half-life (Figure 16). Indoor-resting exposure is derived from the shared vector-behavioural layer, $\text{INDOOR}_r = \text{INDOOR}_n(1 - \rho_{\text{exit}})$, rather than sampled independently, so ITN and IRS exposure assumptions remain biologically coherent.

The combined ITN+IRS mechanism uses sequential exposure. Mosquitoes blocked or killed by a net are not also exposed to IRS during the same feeding attempt, so combined ITN+IRS mortality is slightly below a naive additive formulation at high coverage. In the single-intervention limits, the same mechanism gives the ITN-only and IRS-only effects used in the isolated consistency checks.

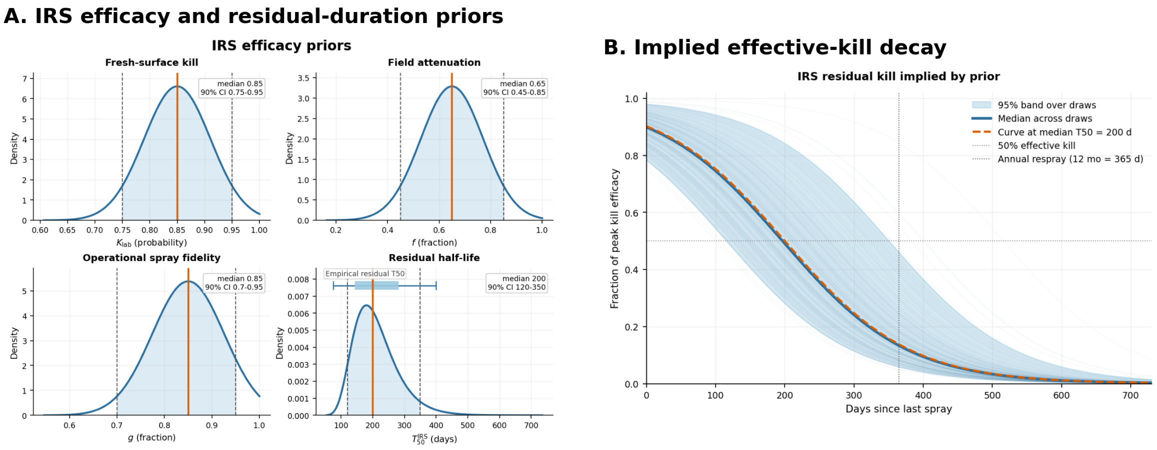


Figure 16: **IRS intervention priors.** Panel A shows fresh-substrate kill, field attenuation, operational spray fidelity, and residual half-life priors. Panel B shows the implied effective-kill decay under annual respraying.

The IRS priors are intentionally generic. Laboratory kill is centered high, field attenuation accounts for incomplete wall coverage, substrate effects, dust, and shorter field contact times, and operational fidelity captures missed structures, refusals, and access barriers within a targeted community. Residual activity is represented by a mixed-substrate half-life prior spanning shorter and longer residual products. The IRS effect-size anchor is correspondingly weaker than the ITN anchor: trial evidence often reflects IRS added to existing ITN coverage or broad Cochrane intervals rather than a clean IRS-vs-control curve. The panel is therefore interpreted as a plausibility check for a generic IRS planning module, not as product-specific validation for pirimiphos-methyl, clothianidin, bendiocarb, or pyrethroid IRS [Tatarsky et al., 2011, Rowland et al., 2013, Ngufor et al., 2017, Sherrard-Smith et al., 2022].

### J SMC priors and effect-size check

Panel C of Figure 14 shows the SMC effect-size consistency check at 80% eligible-child coverage across matched annual EIR, with separate curves for in-season clinical incidence, annual clinical incidence, and annual parasite prevalence. The SMC module uses broad planning priors for prophylaxis half-life, peak per-dose protection, and curative clearance (Figure 17). The four-cycle Jul–Oct schedule and under-five age target follow WHO operational guidance and are treated as scenario controls rather than sampled biological uncertainties.

The prophylaxis half-life prior has median 30 d and a 90% interval of 21–42 d, spanning shorter and longer post-cycle protection profiles in operational and modeling analyses [Cairns et al., 2008, ACCESS-SMC Partnership, 2020]. Peak protection and curative clearance are high immediately after a cycle, but population impact is lower because coverage, adherence, seasonality, and baseline transmission enter downstream. Because SMC trials differ in ITN background, endpoint, seasonality, and reporting window, Panel C uses broad empirical ranges rather than individual EIR-positioned points. The check is separate from the routine-care baseline used in the optimized portfolio figures.

### K Vaccine priors and effect-size check

Panel D of Figure 14 shows the R21 birth-cohort prior-predictive check. In the decision analysis, vaccination reduces force of infection in vaccinated children through a peak protection and exponential waning term (Figure 18). The optimization prior is product-agnostic: peak protection has median $0.75$ with 90% interval $0.55$–$0.90$, and waning half-life has median $540$ d with 90% interval $365$–$730$ d.

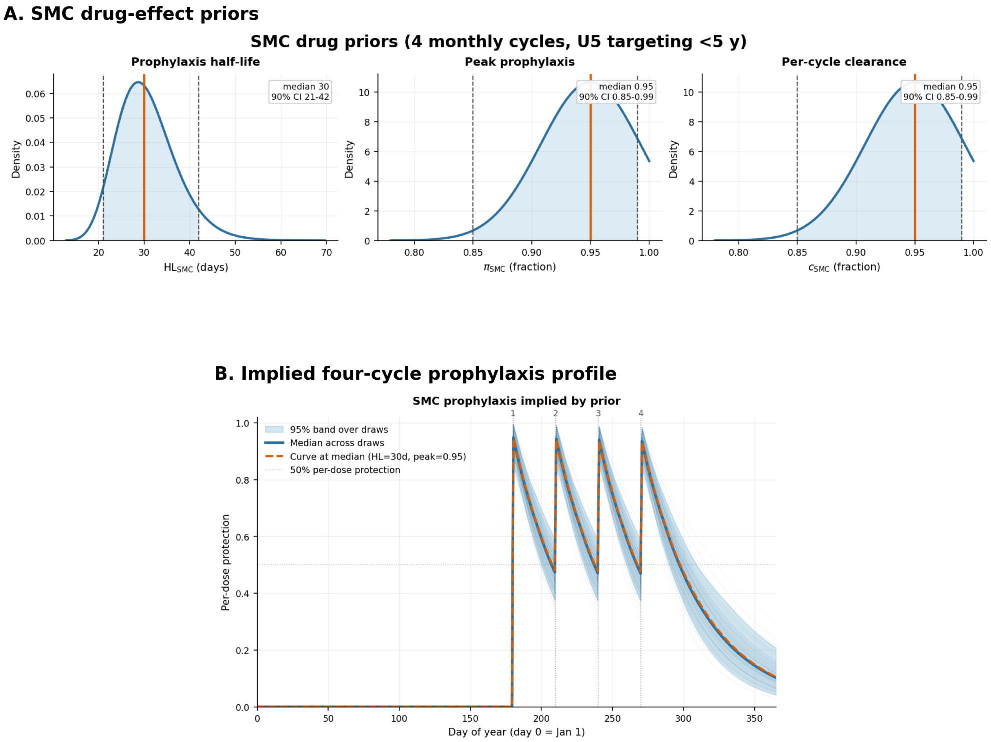


Figure 17: **SMC intervention priors.** Panel A shows the prophylaxis half-life, peak per-dose protection, and per-dose curative-clearance priors. Panel B shows the implied four-cycle Jul–Oct prophylaxis profile.

This deliberately spans RTS,S/AS01 and R21/Matrix-M rather than committing the decision analysis to one product.

**R21 prior-predictive check.** To compare the vaccine priors with R21 trial results, we simulate matched vaccinated and control birth cohorts through the primary 12-month endpoint. The two arms share the same transmission setting, seasonality, and trial-style case management; the vaccinated arm differs only by the waning vaccine protection term. The plotted quantity is $\mathrm{VE} = 1 - \mathrm{cum}_{\mathrm{vacc}}/\mathrm{cum}_{\mathrm{control}}$.

R21 Phase 3 sites differ in transmission intensity, standard of care, and seasonality [Datoo et al., 2024], so Panel D plots vaccine efficacy against each site's reported control-arm clinical incidence rather than assigning sites to nominal model EIR values. The R21 check uses a higher product-specific peak-protection prior (median 0.95 with 90% interval 0.85–0.99) while sharing the generic waning prior. On this axis, the prior-predictive bands overlap the trial confidence intervals across the Phase 3 range. The residual pattern is a somewhat steeper decline in modeled efficacy at high control incidence, so the panel should be read as compatibility evidence for the vaccine module rather than product-specific vaccine calibration.

#### L Robustness of the optimized portfolios to intervention efficacy misspecification

The intervention efficacy and durability parameters described in the main text are planning-scale literature priors that are forward-propagated but *not* updated by the eight-site calibration. The prior-predictive checks above ask whether those priors produce plausible effect sizes. Here we ask a decision-specific question: if an efficacy prior were mis-centred, would deploying the nominal optimized portfolio sacrifice meaningful impact?

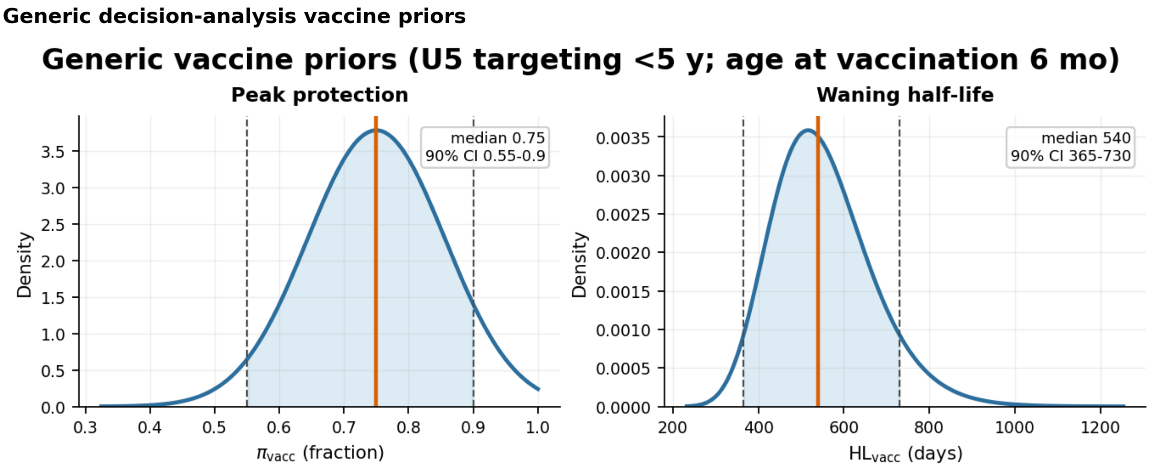


Figure 18: **Generic vaccine priors.** The figure shows the product-agnostic decision-analysis priors for peak infection-blocking protection and exponential waning half-life. These priors deliberately span RTS,S/AS01 and R21/Matrix-M rather than committing the decision analysis to one product.

**Regret metric.** For each perturbation, we shift an efficacy prior centre while preserving its sampled spread, re-optimize the portfolio in the perturbed world, and compare it with the nominal-efficacy optimum deployed in that same perturbed world. Deployment regret is

$$R = \mathcal{U}(\theta_p^\star, p) - \mathcal{U}(\theta_0^\star, p),$$

reported as a percentage of the perturbed-world optimum. A cell is material if regret exceeds $5\%$. We evaluate 18 cells per scenario: EIR $\{3, 10, 30\}$, budgets \$2, \$7, and \$15 per resident over three years, and both under-five and all-age objectives.

**Stress scenarios.** The battery includes adverse one-at-a-time shifts for ITN kill, IRS field efficacy, SMC peak protection and half-life, and vaccine peak protection and waning; joint vector-control, biomedical, and all-intervention adverse scenarios; a favourable high-peak/fast-waning vaccine scenario; a high-EIR faster-waning vaccine scenario; and two below-prior stress probes for IRS and SMC. Table 5 reports the maximum regret for each scenario.

**Result.** Across all $11 \times 18 = 198$ cells under the routine-care baseline, no cell exceeds the $5\%$ materiality threshold. The worst single-cell regret is $2.77\%$, the all-four-adverse scenario reaches $1.98\%$, and below-prior stress probes reach at most $0.79\%$. Median regret is below $0.1\%$. Thus, within this stress-test battery, intervention effect-size priors can be substantially mis-centred without changing the actionable portfolio decision: the uncertainty changes predicted impact more than it changes the optimized allocation.

#### M Garki Project calibration extension

The eight-site calibration includes three pre-intervention villages from the Garki Project (Nigeria, 1970–1972) [Molineaux and Gramiccia, 1980]: **Matsari**, **Rafin Marke**, and **Sugungum**, chosen to span the transmission gradient of the Sudan savanna study area (Sugungum highest, Rafin Marke lowest). Relative to the five cross-sectional sites, Garki adds seasonally resolved surveys and gametocyte-based infectiousness targets.

##### Data aggregation

For each Garki village, we aggregate pre-intervention surveys into age-bin by survey cells using records with completed schedules and collected blood slides before April 1, 1972. Each cell contributes two likelihood targets: standard-microscopy-equivalent parasite prevalence and gametocyte-derived expected human-to-mosquito infectiousness.

Table 5: **Deployment regret under efficacy misspecification.** Maximum regret (percent of the perturbed-world optimum) over the 18 EIR–budget–objective cells per scenario; a material cell exceeds $5\%$. Zero of 198 cells are material; worst single cell $2.77\%$.

| Scenario | Type | Material cells | Max regret |
|---|---|---|---|
| ITN kill $0.40 \to 0.18$ | adverse | 0/18 | $0.09\%$ |
| IRS field $0.65 \to 0.49$ | adverse | 0/18 | $0.51\%$ |
| SMC peak $0.80$, half-life 21 d | adverse | 0/18 | $0.11\%$ |
| Vaccine peak $0.55$, half-life 365 d | adverse | 0/18 | $2.77\%$ |
| ITN + IRS adverse | joint | 0/18 | $0.61\%$ |
| SMC + vaccine adverse | joint | 0/18 | $2.57\%$ |
| All four adverse | joint | 0/18 | $1.98\%$ |
| Vaccine peak $0.90$, half-life 400 d | favourable | 0/18 | $0.05\%$ |
| Vaccine faster-waning at high EIR | context | 0/18 | $0.48\%$ |
| IRS field $0.42$ (below prior) | stress | 0/18 | $0.79\%$ |
| SMC peak $0.55$ (below prior) | stress | 0/18 | $0.13\%$ |

Prevalence targets use the microscopy-sensitivity correction described below. Infectiousness targets are constructed by mapping binned gametocyte densities to mosquito infection probability with the Churcher et al. 2013 saturating dose-response [Churcher et al., 2013]; the cell target is the frequency-weighted average across density bins. This yields roughly 200 calibration cells across five age bins, up to seven pre-intervention surveys per village, three villages, and two observables.

The resulting Garki targets are shown together with the model fit in the main-text posterior-predictive calibration figure. The microscopy correction used to place Garki prevalence on the same observation scale is shown below.

##### EIR forcing strategy

Published annual EIR estimates for these three villages differ across the original monograph and later model-based analyses. We use the monograph values as prior centres, while allowing each village's annual EIR scalar to be learned jointly with the other calibrated parameters (Table 6).

Table 6: Garki annual EIR context and priors. Published annual EIR estimates are shown for context; the calibration estimates its own annual scalar $\kappa_s$ using the monograph value as the log-normal prior centre with Garki-specific width $\sigma_{\log} = 0.28$.

| Site | Monograph | OpenMalaria | EMOD seasonal | $\sigma_{\log}$ | Approx. 95% prior interval |
|---|---|---|---|---|---|
| Matsari | 68 | 68 | 129 | 0.28 | $[39, 118]$ |
| Rafin Marke | 18 | 18 | 39 | 0.28 | $[10, 31]$ |
| Sugungum | 145 | 132 | 80 | 0.28 | $[84, 251]$ |

The OpenMalaria estimates are from Reiker et al. [2021]; the EMOD seasonality calibration estimates are from Selvaraj et al. [2018]. Given the dispersion among published estimates, we treat each site's annual EIR as a calibratable parameter rather than fixing it to one literature value. The monthly forcing is constructed as

$$\mathrm{EIR}^{(s)}_{\text{monthly}}(m) = \kappa_s \cdot \tilde{\phi}_s(m),$$

where $\kappa_s$ is the calibrated annual-EIR scalar and $\tilde{\phi}_s$ is the normalised monthly seasonal shape (summing to 1) from Selvaraj et al. [2018]. Those shapes provide a smooth village-specific seasonal profile, including low but non-zero dry-season forcing.

##### What Garki adds to the calibration

Garki adds information that the five cross-sectional sites do not: seasonal age-prevalence trajectories, gametocyte-derived human-to-mosquito infectiousness targets, and village-specific EIR scalars estimated jointly with the rest of the posterior. These features help calibrate the infectiousness kernel and test whether the model can reproduce wet- and dry-season age patterns, rather than only equilibrium snapshots.

##### Observation-process correction: Garki microscopy $\rightarrow$ standard-microscopy-equivalent prevalence

The Garki microscopy protocol examined more blood per slide than the standard microscopy and RDT endpoints used in the other calibration sites, making it more sensitive to low-density chronic infections. Raw Garki microscopy prevalence is therefore not directly comparable to the other prevalence targets, especially in adults.

To put Garki on the same observation scale, we convert each individual Garki microscopy record to a predicted standard-microscopy-equivalent result using a parasite-density posterior and a standard-microscopy sensitivity curve, then aggregate to the site–survey–age cells used in the likelihood. The correction is strongly age-graded: the median standard-equivalent/raw prevalence ratio is about 0.70 in children under 5, 0.58 at ages 5–15, and 0.23–0.24 in adults. After correction, the Garki adult age-prevalence curves are qualitatively comparable to the other high-transmission calibration sites while preserving the within-site seasonal signal (Figure 19).

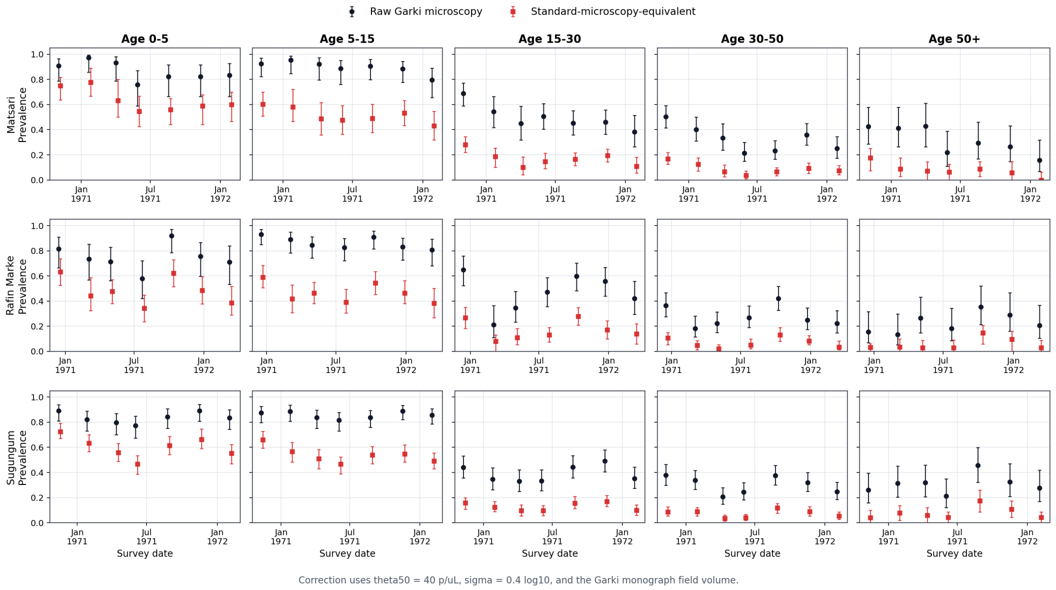


Figure 19: Raw Garki microscopy prevalence (black) vs. standard-microscopy-equivalent prevalence (red) across all (site, survey, age) cells. The correction lowers adult prevalence while preserving the within-site seasonal signal.

The corrected prevalence targets replace raw microscopy prevalence in the Garki likelihood; the gametocyte-derived infectiousness targets are unchanged. This is a preprocessing correction only: the compartmental model is unmodified, and no Garki-specific detection parameters are introduced.

#### N Posterior estimation and NUTS diagnostics

This appendix documents the No-U-Turn Sampler (NUTS; Hoffman and Gelman 2014) run that serves as the posterior of record. SVI was used only as computational scaffolding for initialization and preconditioning; all posterior summaries in the manuscript use the retained four-chain NUTS sample.

##### Posterior target

The NUTS chain targets the identical NumPyro model used by SVI: the same combined likelihood, bounded-parameter transform, and prior terms define both targets. For each non-Garki calibration site $s$ and age bin $a$ with observed diagnostic positives $x_{s,a}$ out of $n_{s,a}$ sampled, the prevalence term is

$$x_{s,a} \sim \text{BetaBinomial}(n_{s,a},\, \mu_{s,a}(\theta),\, \phi)\,, \qquad \phi = 1/\rho_{\text{prev}} - 1,$$

where $\mu_{s,a}(\theta)$ is the model-predicted diagnostic positivity and $\rho_{\text{prev}} = 0.02$ in the production fit. For clinical incidence counts $c_{s,a}$ over person-years $\text{PY}_{s,a}$,

$$c_{s,a} \sim \text{NegBin}\big(\text{PY}_{s,a}\, \lambda^{c}_{s,a}(\theta),\, r_{\text{inc}}\big)\,, \qquad r_{\text{inc}} = 1/\text{CV}^2_{\text{inc}},$$

with $\text{CV}_{\text{inc}} = 0.5$. Sites lacking sample-size metadata use $n_{\text{eff}} = 50$ per age bin. For the three Garki villages, the same beta-binomial prevalence likelihood is evaluated at each pre-intervention survey month and age bin, while the reconstructed human-to-vector infectiousness target $\bar{c}_{s,t,a}$ is modeled with a beta likelihood centered on the model-predicted $\bar{c}(\theta)$ and cell-specific concentration parameters derived from the infectiousness bootstrap intervals, capped at 100. The 38 bounded calibrated parameters are represented in unconstrained coordinates by

$$x_j(z_j) = \ell_j + (u_j - \ell_j)\sigma(z_j),$$

where $\ell_j$ and $u_j$ are the parameter bounds and $\sigma$ is the logistic function. The NUTS log density in $z$ is

$$\log p(z \mid \mathcal{D}) = -\mathcal{L}_{\text{combined}}\{x(z)\} + \sum_j \log \left| \frac{dx_j}{dz_j} \right| + \text{const}.$$

The second term is the sigmoid change-of-variables log-Jacobian. Its inclusion is essential: it makes the unconstrained $z$ target exactly the posterior induced by the bounded-parameter density, rather than an improper target with flat tails in weakly identified directions.

##### NUTS run and diagnostics

NUTS was initialized from the eight-site `AutoMultivariateNormal` SVI guide and used the fitted full-rank SVI covariance as a fixed dense metric. This affects sampler efficiency, not the stationary posterior target. The production run used four vectorized chains, target acceptance probability 0.90, and maximum tree depth 9. The retained posterior contains 540 post-warmup draws per chain, or 2160 pooled draws.

The completed run passed standard HMC and chain-mixing diagnostics [Vehtari et al., 2021]: zero divergent transitions, no tree-depth saturation (mean 5.2, maximum 8), split-$\hat{R} < 1.05$ for all 38 calibrated parameters, and median effective sample size $\approx 1{,}700$ (Figure 20). The one slow-mixing parameter is the weakly identified infectiousness coefficient $a_{m,\text{inf}}$ (ESS $\approx 75$), whose marginal is broad but has little posterior-predictive influence. NUTS and the SVI guide occupy the same likelihood-supported region, while NUTS produces wider marginal intervals (median NUTS/SVI 95% width ratio 1.35, never below one; Figure 21). We therefore use NUTS as the posterior of record and SVI as a rapid development and preconditioning tool.

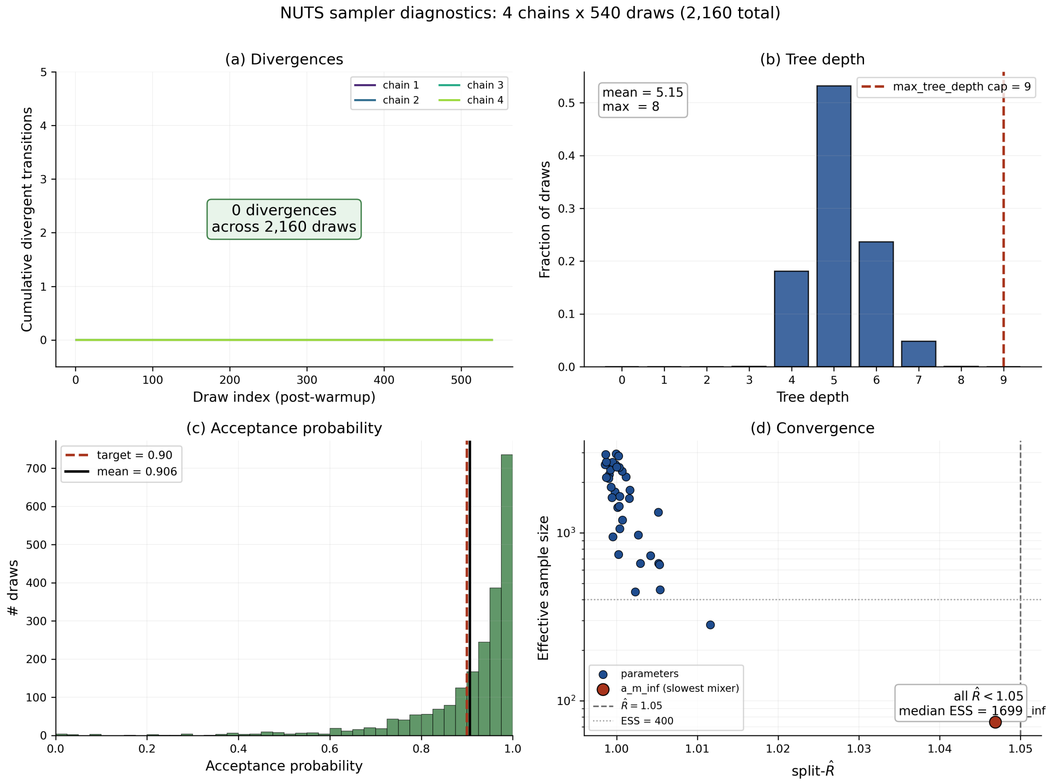


Figure 20: **NUTS sampler diagnostics.** (a) Cumulative divergent transitions by draw for each of the four chains: zero across all retained draws. (b) Distribution of per-draw tree depth, well below the `max_tree_depth` cap of 9 (mean 5.2, maximum 8). (c) Distribution of realized acceptance probability (mean 0.91; target 0.90). (d) Per-parameter effective sample size versus rank-normalized split-$\hat{R}$: all 38 parameters satisfy $\hat{R} < 1.05$ with median ESS $\approx$ 1,700, the lone slow mixer being the weakly identified $a_{m,\mathrm{inf}}$.

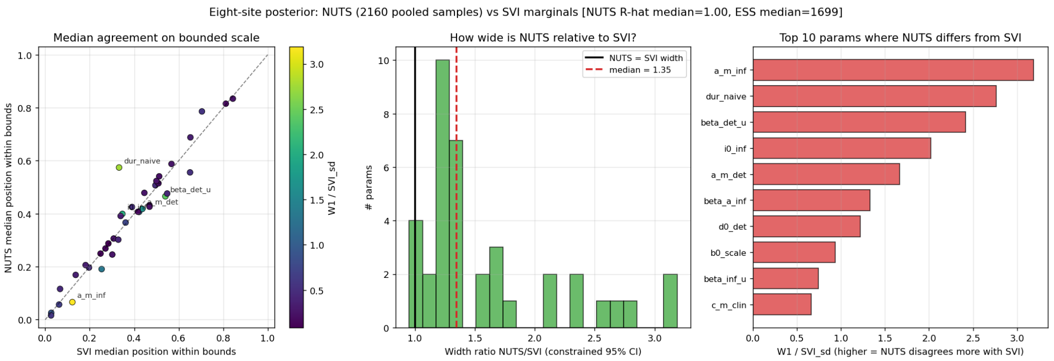


Figure 21: **Parameter-space comparison of SVI and NUTS.** Left: NUTS versus SVI posterior medians for all 38 parameters on their bounded $[0, 1]$ scale, colored by 1-Wasserstein distance in SVI standard-deviation units; the medians agree closely (median shift 0.25 SD). Center: distribution of the NUTS/SVI 95% credible-interval width ratio (median 1.35); the ratio never falls below one, so the Gaussian SVI guide never overstates uncertainty. Right: the ten parameters where NUTS departs most from SVI, led by the weakly identified $a_{m,\mathrm{inf}}$ and $\mathrm{dur}_{\mathrm{naive}}$.

World Health Organization. WHO recommends R21/Matrix-M vaccine for malaria prevention in updated advice on immunization, October 2023. News release; reports R21 price range of US$2–US$4 per dose and notes RTS,S/R21 product choice should depend on programme characteristics, supply, and affordability.

Joshua Yukich, Peder Digre, Sara Scates, Luc Boydens, Emmanuel Obi, Nicky Moran, Allison Belemvire, Mariandrea Chamorro, Benjamin Johns, Keziah L. Malm, et al. Incremental cost and cost-effectiveness of the addition of indoor residual spraying with pirimiphos-methyl in sub-Saharan Africa versus standard malaria control: results of data collection and analysis in the Next Generation Indoor Residual Sprays (NgenIRS) project, an economic-evaluation. *Malaria Journal*, 21(1):185, 2022. doi: 10.1186/s12936-022-04160-3.

Joshua O Yukich, Christian Lengeler, Fabrizio Tediosi, Nick Brown, Jo-Ann Mulligan, Des Chavasse, Warren Stevens, John Justino, Lesong Conteh, Rajendra Maharaj, et al. Costs and consequences of large-scale vector control for malaria. *Malaria Journal*, 7(1):258, 2008. doi: 10.1186/1475-2875-7-258.
